# Computational Linguistic Alignment in Psychosis from Semi-Structured Clinical Interviews

**DOI:** 10.64898/2026.05.24.26353973

**Authors:** Emmanuel Olarewaju, Alban Voppel, Fiona Meister, Chaimaa El Mouslih, Paulina Dzialoszynski, Lena Palaniyappan

**Affiliations:** Department of Psychiatry, McGill University, Montréal, Quebec, Canada; Douglas Mental Health University Institute, Montréal, Quebec, Canada; Integrated Program in Neuroscience, McGill University, Montréal, Quebec, Canada; Department of Psychology, McGill University, Montréal, Quebec, Canada; Robarts Research Institute, Western University, London, Ontario, Canada; Department of Medical Biophysics, Western University, London, Ontario, Canada; Montreal Bilingualism Initiative, McGill University, Montréal, Quebec, Canada; Center for Research on Brain, Language and Music, McGill University, Montréal, Quebec, Canada

**Author notes:** **Corresponding author** Lena Palaniyappan.

**Keywords:** Conversational alignment, Schizophrenia spectrum disorder, Formal thought disorder, Psycholinguistics, Dyadic interaction, Sentence embeddings

## Abstract

**Background:** Something in discourse with a person experiencing psychosis often “feels off” before formal assessment is completed, yet this phenomenon has not been quantified at the level of ongoing dyadic conversation. Prior work has largely treated patient speech in isolation, limiting our capacity to measure how communicative disruption emerges within clinical exchange.

**Methods:** We applied a three-level decomposition of conversational alignment in 109 patients with psychotic disorders (26 female) and 60 healthy controls (22 female) at baseline and 12 months (*n* = 115). Register divergence captured lexical distance between interviewer and participant; embedding-based synchrony measured semantic trajectory coupling; within-speaker coherence was computed separately for each speaker. We used linear mixed-effects models adjusted for timepoint and participant clustering.

**Results:** Patients showed significantly greater lexical-semantic divergence from the interviewer (*d* = 0.48, *p* < .001) and reduced embedding-based synchrony (*d* = −0.59, *p* < .001), both effects observed at each timepoint, consistent with a partially stable profile. Critically, the interviewer’s within-speaker coherence was reduced during conversations with patients (d = −0.33, p = .016), indicating that the linguistic phenomenon of interest extended beyond patient speech to the interaction itself. Register divergence tracked impoverished thinking, while synchrony tracked disorganized thinking (both FDR-corrected *q* = .038).

**Conclusions:** Conversational alignment in psychosis reveals a reconfiguration of dyadic semantic coordination, marked by reduced interviewer coherence despite preserved patient narrative continuity. These transcript-derived alignment metrics offer a scalable approach to quantifying interpersonal communicative function in routine clinical encounters.

## 1. Introduction

Clinical interviews with patients with schizophrenia can feel anomalous before formal assessment. Rümke called this the *Praecox Feeling* ^(1)^ with a claim that “schizophrenia is not diagnosed by examining the patient, but by examining one’s own inner world” (also see Minkowski’s statement “in my contact with the patient, something broke within me” ^2^). Modern phenomenologists have described this experience as a difficulty in ‘attunement’ that distinguishes schizophrenia from other psychoses with replicable discrimination ^(3–6)^. The phenomenological focus on meaning and rhythm raises the question of whether speakers inhabit a shared communicative space, using similar words and levels of expression ^(7–10)^.

Two approaches have attempted to quantify speech in schizophrenia: psychopathological rating scales for formal thought disorder (FTD; ^11–16^) and language-focused research (^17–26^). Both describe the structure of patient speech in isolation, overlooking how a patient influences the other speaker. Impaired conversational pragmatics in schizophrenia spectrum disorder ^(27–29)^ strongly influence social and occupational functioning ^(30–32)^, but the interactional component of this breakdown remains largely unmeasured, with recent work beginning to quantify this domain as linguistic synchrony between speakers and linking it to real-world social functioning in early psychosis ^(33)^. Cooperative communication depends on speakers jointly scaffolding meaning across turns, shaping each contribution so that it is informative, contextually relevant, and interpretable without excessive inferential burden for the conversational partner ^(34)^. This process is highly impaired in schizophrenia but remains understudied ^(29)^. Here we focus on this alignment, situating it within a broader literature demonstrating that interpersonal attunement can be quantified across multiple levels, from controlled paradigms to naturalistic conversation.

Dialogue is organized through a dyadic relation between speaker and interlocutor ^(35)^. Treating the patient’s transcript alone as the interview readout discards interpersonal information. Interactive alignment ^(36,37)^ (recently invoked in describing formal thought disorder in schizophrenia ^38^) specifies what is being discarded when we restrict focus to one side of a dialogue. In naturalistic interactions, speakers prime each other across phonological, lexical, syntactic, and semantic strata in real time. This process generates progressive convergence of linguistic representations across speakers, establishing a “common ground” for continued dialogue. Restricting analysis to a single speaker removes access to whether conversational partners are tracking each other’s register (i.e., the level and structure with which language is expressed) and moving through meaning together. The degree of alignment provides an index of the precision and coordination of this dyadic system. Accordingly, in clinical settings, the interviewer is not a passive contributor but an active co-constructor of the exchange.

When studying cooperative dynamics, most recent work has focused on paralinguistics, e.g., temporal and acoustic properties of turn-taking, with reduced turn length and a greater need for repeated prompts (higher turn frequency) associated with the severity of negative symptoms in schizophrenia (^39–43^; also see ^44^ for a broader review). Qualitative work on therapeutic alliance formation similarly demonstrates that clinical outcomes depend on mutual engagement, rupture, and repair unfolding across early interactions, rather than properties of either participant alone ^(45)^.

Relatively few studies have directly examined how meaning at the lexico-semantic level is aligned between patients and interviewers (e.g., Achim et al., ^46^; Sharpe et al., ^47^; see also Dwyer et al., ^48^). Dwyer and colleagues ^(48)^ demonstrated reduced lexico-semantic alignment in an interactive maze game, but this word-level misalignment was not observed when a picture-naming game was used ^(47)^. These task paradigms operate under controlled conditions that do not capture the open-ended, temporally evolving dynamics of clinical exchange. How linguistic alignment evolves during conversation, persists over time, and affects the interviewer’s contribution remains unclear. Addressing these questions enables the transition from single-speaker descriptions to time-resolved, dyadic measures of interaction, providing a behavioural readout of real-time coordination between people experiencing schizophrenia and their clinical interlocutors ^(32,49)^.

We propose that conversational alignment can be decomposed into three conceptually distinct levels: register alignment, within-speaker coherence, and synchrony (see Table 1).

**Table 1.**
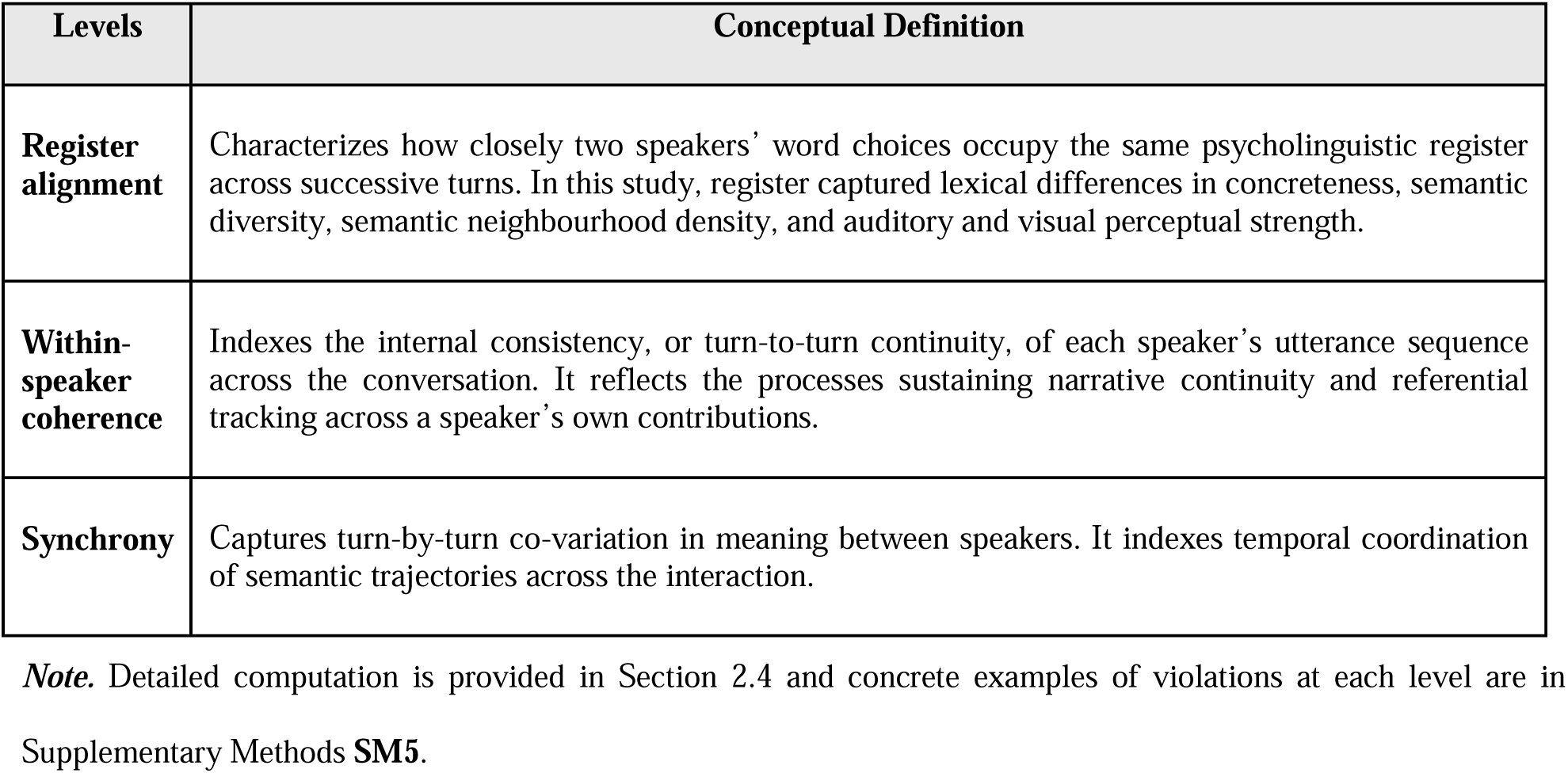
Three conceptual levels of conversational alignment.

These three levels are operationally as well as mechanistically distinct. A patient may converse from a divergent register while moving in tandem, unconscious (speech and bodily) alignment with the interviewer ^(50)^, without affecting the interviewer’s own semantic coherence. Decomposing alignment at these three levels makes it possible to distinguish, within a single transcript, which aspect of communicative function is preserved, and which is altered.

We tested four questions. First, do register divergence, synchrony, and within-speaker coherence distinguish patients from healthy controls as separable dimensions of communicative dysfunction (*diagnostic effect*)? Second, do these alignment metrics map onto distinct clinical features, including impoverished thinking, disorganized thinking, and functional capacity, or do they reflect a single undifferentiated process (*symptom relevance)?* Third, do these metrics show longitudinal persistence and covary with within-person clinical change over 12 months (*state vs. trait-like effect*)? Fourth, how does alignment vary across the semi-structured order of the interview, and where are patient–control differences most evident (*context effect*)?

## 2. Methods

### 2.1. Participants

Patients were recruited through the Prevention and Early Intervention Program for Psychoses (PEPP) at London Health Sciences Centre, a defined catchment-area programme serving first-episode and recent-onset psychosis in Southwestern Ontario, Canada. Healthy controls (HC) were recruited from the same geographic catchment area via community advertisement. Eligibility for the patient group required meeting DSM-5 criteria for a psychotic disorder, made by a staff psychiatrist and confirmed via Leckman’s ^(51)^ best estimate consensus procedure. Participants with psychosis secondary to a substance use disorder, an active substance dependence in the past year, or a known neurological disorder affecting speech output (e.g., apraxia) were excluded. Eligibility for healthy controls required no personal or first-degree family history of psychosis.

At baseline, 169 participants were enrolled: 109 patients (PT; 83 male, 26 female; mean age 28.6 ± 7.6 years) and 60 healthy controls (HC; 38 male, 22 female; mean age 28.2 ± 7.8 years). Groups did not differ significantly in age (*t*-test, *p* > .05). Patient diagnoses (consensus procedure): schizophrenia *n* = 58, schizoaffective disorder *n* = 24, psychosis not otherwise specified *n* = 22, bipolar disorder with psychotic features *n* = 5. A 12-month follow-up assessment was completed by 70 of 109 patients and 45 of 60 HC. Participants with baseline enrolment data but no baseline interview transcript (n=2) were excluded from all baseline and change-score analyses (see Sankey figure **SM2**). The final analytic samples after removing incomplete audio/transcripts: Baseline, PT *n* = 99 transcripts, HC *n* = 57 transcripts; 12-month, PT *n* = 70 transcripts, HC *n* = 45 transcripts. Written informed consent was obtained from all participants. All procedures were approved by the Research Ethics Board at Western University (London, Ontario). Part of this cohort has been analyzed in a complementary single-speaker study ^(52)^; the present dyadic alignment analyses do not overlap with that report.

### 2.2. Interview Procedure

Trained interviewers conducted all interviews at baseline and at 12-month follow-up using the DISCOURSE semi-structured protocol (discourseinpsychosis.org). The protocol comprised seven sequential discourse contexts: free conversational speech, personal narrative, health narrative, picture description, storyboard narration, dream report, and reading and recall. These contexts varied in cognitive demand, external scaffolding, and opportunity for reciprocal exchange. To ensure comparability, the section order, instructions, and 14 initiating interviewer prompts/questions were standardized. Beyond these predefined elements, speech and contingent exchanges were unscripted and unfolded naturally, except during the prescribed reading-aloud component. Across sections, interviewers could rephrase questions and provide non-directive prompts while otherwise minimizing verbal intervention. Interviews lasted 15–35 minutes.

### 2.3. Data Collection and Transcription

Audio was recorded with a digital voice recorder (Olympus/SONY) positioned centrally on the assessment table less than four feet from both speakers. Transcriptions were generated automatically from audio using *Batchalign* (a Python program producing CHAT-format transcripts via the TalkBank/CLAN system ^53^), then manually verified by qualified researchers against the original recordings. Verified transcripts were stored on the PsychosisBank (https://talkbank.org/access/English/Palaniyappan/Discourse-UWO.html) and converted to plain-text format with speaker prefix labels (Interviewer / Participant) for computational analysis. The full corpus comprised 273 transcripts across both timepoints.

### 2.4. Computational Alignment Pipeline

All alignment metrics were computed using a custom Python pipeline applied to the plain-text transcripts. Each transcript was parsed into an Interviewer (IV) turn sequence and a Participant (PT) turn sequence by merging consecutive same-speaker lines. Two parallel feature systems were used: psycholinguistic norm composites and MiniLM sentence embeddings ^(54)^.

For the psycholinguistic composites, content words in each turn were extracted by lemmatization and stop word removal (NLTK WordNetLemmatizer ^55^; combined stop word list). Each content word was scored on five psycholinguistic dimensions derived from the ConversationAlign framework ^(56)^: a) Concreteness: rated 1–5 from ^(57)^. Higher values indicate more sensory-perceptually grounded vocabulary. b) Semantic diversity: from ^(58)^; indexes contextual promiscuity i.e., how consistently a word appears across similar versus varied discourse contexts. c) Semantic neighbourhood density: log co-occurrence count from ^(59)^; indexes position in the co-occurrence structure of the lexicon. d) Visual perceptual strength and e) auditory perceptual strength from ^(60)^; degree to which word meaning engages the visual or the auditory modality. All five dimensions were rescaled to a common [0, 1] range prior to aggregation to derive a per-turn norm composite (mean across the five-dimension means, requiring at least three non-missing dimensions for a turn to be included). Participants with fewer than eight valid turns were excluded from the full pipeline. The norm composite produced a scalar time series per speaker per conversation: *iv_norm* and *pt_norm*.

For the sentence embeddings, each turn (full merged text) was encoded by *paraphrase-multilingual-MiniLM-L12-v2* (*SentenceTransformers;* ^61^), *producing a 384-dimensional dense vector*. All interviewer and participant turns within a conversation were encoded, producing *iv_embeds* and *pt_embeds* as arrays of shape *n* × 384.

Three primary metrics were computed for each conversation at each timepoint, each capturing a distinct dimension of the alignment framework; see Table 2 for metrics. A summary of metric units and illustrative register examples is provided in Supplementary Methods **SM5**.

**Table 2.** Primary conversational alignment metrics.

| Metric | Feature system | Computation | Unit of analysis | Higher values indicate |
| --- | --- | --- | --- | --- |
| **1. Register divergence, $dAUC_{norm}$** | Psycholinguistic norm composites | Absolute interviewer–participant norm differences were integrated across paired turns: $dAUC_{norm} = \text{trapz}(\vert iv\_norm[t] - pt\_norm[t]\vert ) / (n\_pairs - 1)$, where $n\_pairs$ is the number of paired turns. | Conversation/timepoint | Greater register divergence between interviewer and participant. |
| **2. Within-speaker coherence** | Sentence embeddings | Cosine similarity was computed between consecutive turn embeddings separately for IV and PT:<br>$iv\_cont[t] = \cos(iv\_embeds[t-1], iv\_embeds[t])$;<br>$pt\_cont[t] = \cos(pt\_embeds[t-1], pt\_embeds[t])$, for $t = 2..n$. | Speaker/conversation/timepoint | Greater turn-to-turn continuity within a speaker's own contributions. |
| **3. Embedding-based synchrony, $r_{\text{embed}}$** | Sentence embeddings | IV and PT within-speaker continuity streams were calculated as consecutive-turn cosine similarities and aligned by paired exchange index after same-speaker lines were merged. Streams were truncated to the minimum shared length. $r_{\text{embed}}$ was computed as the Pearson correlation between the aligned continuity streams. | Conversation/timepoint | Stronger turn-by-turn semantic coupling between speakers. |

**Note.** IV = interviewer; PT = participant; $\text{dAUC}_{\text{norm}}$ = norm-based divergence area under the curve; $\text{trapz}$ = trapezoidal estimate of area under the turn-by-turn difference curve; $r_{\text{embed}}$ = embedding-based synchrony; $n_{\text{pairs}}$ = number of paired interviewer-participant turns. Metrics were computed for each conversation at each time point. Within-speaker coherence was computed separately for each speaker.

For comparison with register divergence (dAUCnorm), we computed divergence over turn embeddings (dAUCembed). A joint PCA was fitted to all interviewer and participant turn embeddings within each conversation, with both speakers’ turns stacked before decomposition. Only the first principal component (PC1) was retained, capturing the dominant semantic axis of within-conversation variation. Projecting each turn embedding onto PC1 produced separate scalar trajectories for each speaker. These trajectories were aligned by paired exchange index, and divergence was computed as the normalized area under the absolute interviewer-participant difference curve: dAUC_embed_ = trapz(|iv_pc1[t] − pt_pc1[t]|) / (n_pairs − 1). A component-retention sensitivity analysis (1–10 components) is reported in Supplementary Results SR13.

**Figure 1.**
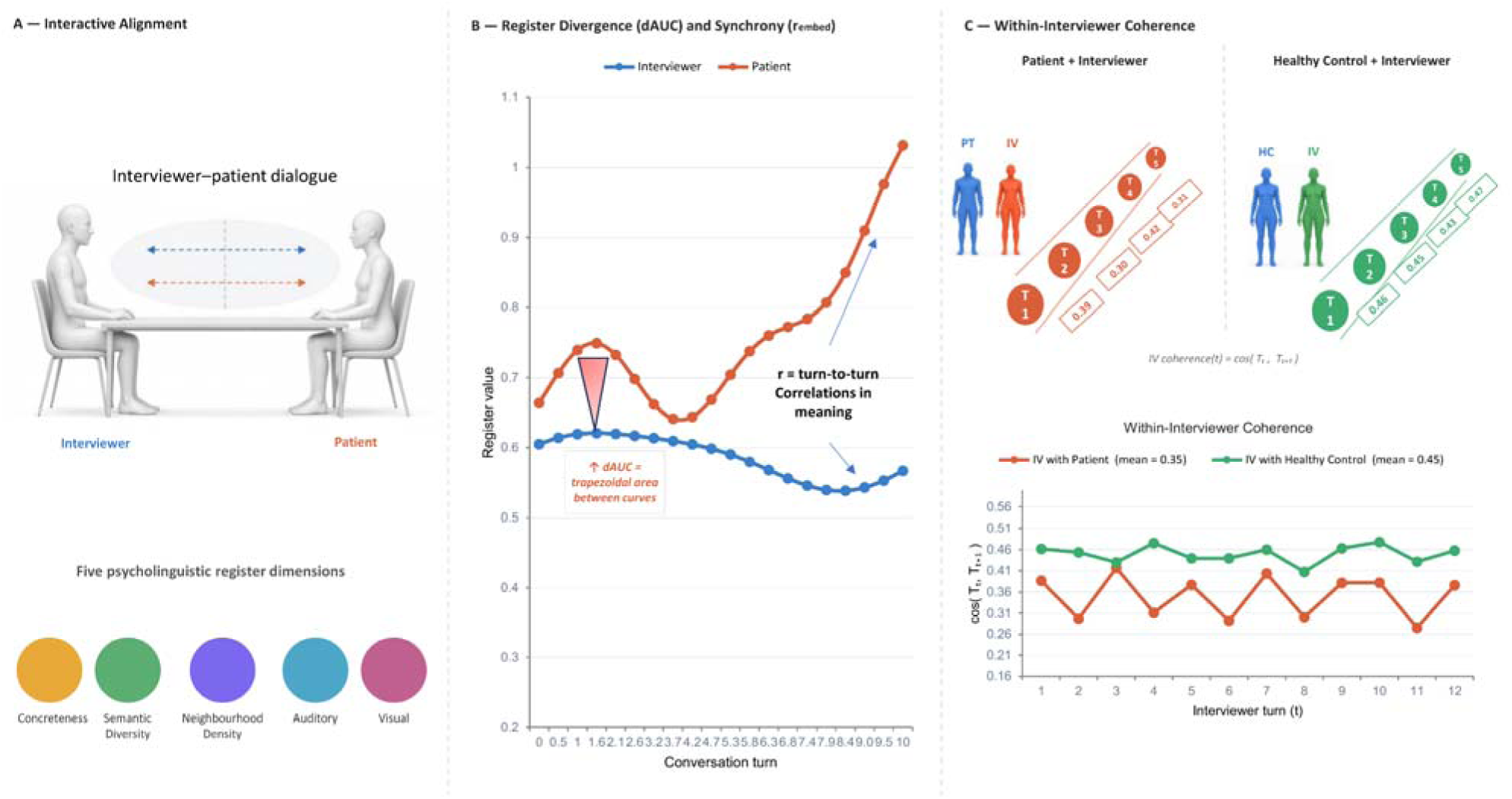
Conceptual framework for the study of conversational alignment. Schematic illustration of the three-level decomposition of conversational alignment applied in the present study. <u>Left</u>: Conversational registers and synchrony of navigation through semantic space are captured by studying dyadic interaction. Register divergence is quantified using the psycholinguistic profile of word choices across five semantic dimensions selected for their relevance to schizophrenia-spectrum language disturbance: concreteness, semantic diversity, neighbourhood density, auditory perceptual strength, and visual perceptual strength (see Supplementary Methods **SM1**). <u>Middle</u>: Divergence is operationalized as the normalized trapezoidal area under the turn-by-turn divergence curve (dAUC_norm_). Embedding-based synchrony captures the degree to which both speakers’ within-speaker semantic continuity streams co-vary turn-by-turn, operationalized as the zero-lag Pearson correlation between consecutive-turn cosine similarity series (r_embed_). <u>Right</u>: Within-speaker coherence captures the internal semantic consistency of each speaker’s own utterance sequence, operationalized as the mean of the within-speaker continuity stream. This is shown for the interviewer in relation to a patient and a healthy participant, respectively, illustrating that interviewer coherence can vary with dyadic context. Representative high and low examples of divergence, synchrony, and coherence are provided in Supplementary Methods **SM5**

### 2.5. Clinical Measures

All clinical assessments were administered at Baseline and at 12-month follow-up by trained research staff. Thought and Language Index (TLI ^62^) was rated based on the picture description task. Two composite scores were computed: *TLI Impoverishment* (Poverty of Speech + Weakening of Goal + Perseveration) and *TLI Disorganization* (Looseness + Peculiar Words + Peculiar Sentences + Peculiar Logic + Distractibility). An abbreviated version of Positive and Negative Syndrome Scale (PANSS ^63^) was completed (P1, P2, P3 summed for positive; N1, N4, N6 for negative symptom scores). Social and Occupational Functioning Assessment Scale (SOFAS ^64^) was also administered at both timepoints. For clinical correlation analyses, all clinical variables were z-scored within the PT sample across both timepoints prior to entry into models. For change-score analyses, variables were additionally standardized within the change-score sample.

### 2.6. Statistical Analysis

#### Group comparisons

All analyses were pre-specified in an OSF Secondary Data Preregistration (https://osf.io/xf8y7). Analyses were conducted in Python 3.10 using statsmodels (version 0.14) MixedLM for all linear mixed-effects models (LMER). Statistical significance threshold was α = .05, two-tailed, throughout unless otherwise specified. Group comparisons pooled Baseline and 12m observations in a single LMER: metric ~ group_bin * tp_c + (1 | participant_ID) where *group_bin* (HC = 0, PT = 1), *tp_c* (Baseline = −0.5, 12m = +0.5; group main effect is estimated at the Baseline-12m midpoint), and *(1 | participant_ID)* is a random intercept per participant accounting for repeated-measures correlation. This model was applied separately to each alignment metric. The primary contrast was the group’s main effect. The Group × Timepoint interaction was also tested. Effect size Cohen’s *d* was computed from LMER beta or partial-r with bootstrap 95% CIs on *d* from 2,000 resamples (percentile method; raw pooled data). Unequal group sizes were accommodated by estimating mixed-effects models on all available observations, with participant-level random intercepts accounting for repeated measures.

To locate the largest PT–HC register gaps, a sliding-window analysis examined per-turn absolute norm divergence across 41 equally spaced window centres (step = 1/40; window width = 0.12 normalized conversation length, approximately 12% of the interview). LMERs were fitted separately within each window, with the group coefficient reflecting the cross-sectional PT–HC gap at each conversation position. Windows with p < .05 for the group effect were identified, and spatially contiguous significant windows were clustered. Task boundaries were identified from the standardized DISCOURSE interviewer prompts and manually verified across transcripts. Speaker-specific directional analyses in embedding space are reported in Supplementary Results SR12 (Figure SF10).

#### Correlational analyses

Associations between alignment metrics and clinical variables were tested using linear mixed-effects models (LMERs) applied to all available observations across baseline and 12-month follow-up. Participants with data at both timepoints contributed two observations, and participant ID was included as a random intercept to account for repeated measurements. Primary clinical-association models adjusted for timepoint and included a participant-level random intercept. An additional group-adjusted sensitivity analysis of TLI Impoverishment is reported in Supplementary Results **SR5**. Four alignment metrics (divergence, *r*_embed_, interviewer and participant coherence) were tested against three clinical variables (TLI Impoverishment, TLI Disorganization, and SOFAS), yielding a 4 × 3 = 12 test matrix.

To examine whether within-person changes in alignment tracked clinical change, change-score analyses were conducted in the paired participant subsample (n = 113). All variables were standardized within the change-score sample and standardized β was reported. Four alignment metrics were related with 3 functional and clinical variables - SOFAS Δ, TLI Impoverishment Δ, TLI Disorganization Δ with Benjamini-Hochberg false-discovery-rate (FDR) correction applied separately to the cross-sectional and change-score tests.

Pairwise alignment-metric associations used the same LMER partial-r methods as clinical correlations, with one metric as outcome and a second as predictor (z-scored), controlling for timepoint and participant-level random effects. Analyses were conducted separately in HC conversations and PT conversations.

## 3. Results

The final analytic sample with intact transcripts comprised 99 patients and 57 healthy controls (HC) at baseline (Baseline) and 70 patients and 45 HC at 12-month follow-up (12m), yielding 271 observations across the two timepoints. For change-score analyses requiring valid transcripts at both assessments, 113 paired participants were available (68 patients [the excluded 2 had no baseline audio, but returned at 12-months], 45 healthy controls); 99 had paired SOFAS scores.

Groups did not differ in age, but patients showed significantly lower educational attainment, lower social and occupational functioning, and higher formal thought disorder scores as expected. Full demographic and clinical characteristics are presented in Table 3.

**Table 3.**
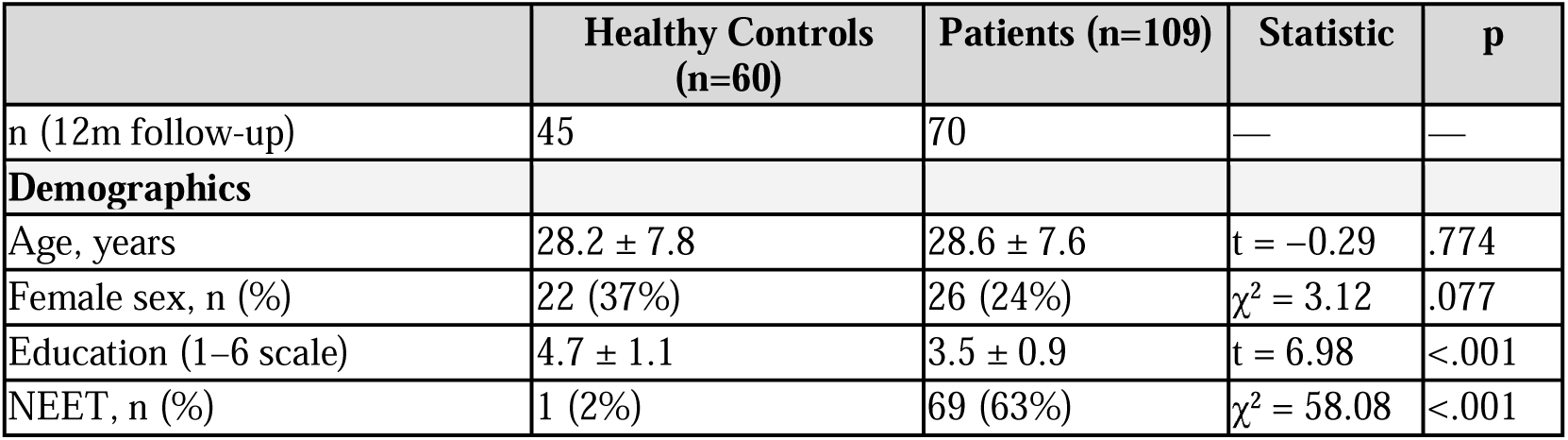
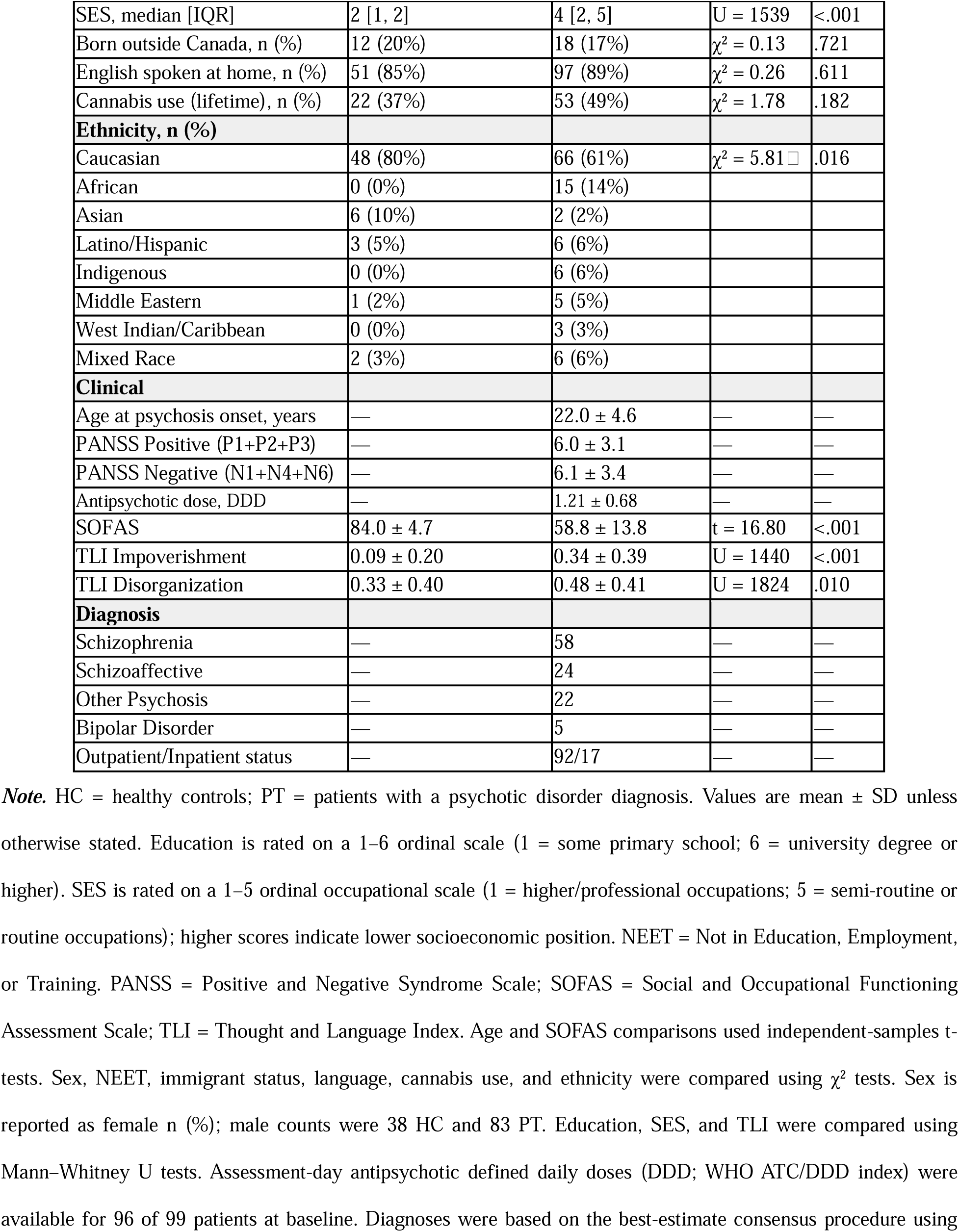
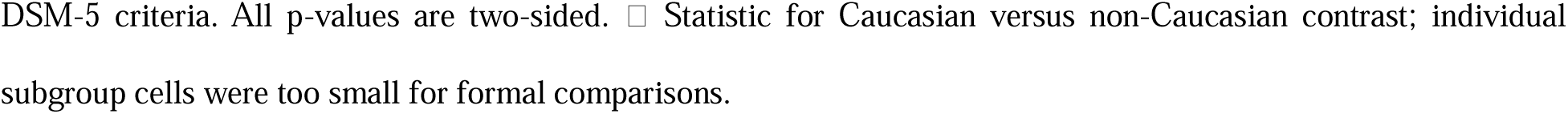
Demographics.

|  | Healthy Controls<br>(n=60) | Patients (n=109) | Statistic | p |
| --- | --- | --- | --- | --- |
| n (12m follow-up) | 45 | 70 | — | — |
| **Demographics** |  |  |  |  |
| Age, years | 28.2 ± 7.8 | 28.6 ± 7.6 | t = -0.29 | .774 |
| Female sex, n (%) | 22 (37%) | 26 (24%) | $\chi^2 = 3.12$ | .077 |
| Education (1–6 scale) | 4.7 ± 1.1 | 3.5 ± 0.9 | t = 6.98 | <.001 |
| NEET, n (%) | 1 (2%) | 69 (63%) | $\chi^2 = 58.08$ | <.001 |
| SES, median [IQR] | 2 [1, 2] | 4 [2, 5] | U = 1539 | <.001 |
| Born outside Canada, n (%) | 12 (20%) | 18 (17%) | $\chi^2 = 0.13$ | .721 |
| English spoken at home, n (%) | 51 (85%) | 97 (89%) | $\chi^2 = 0.26$ | .611 |
| Cannabis use (lifetime), n (%) | 22 (37%) | 53 (49%) | $\chi^2 = 1.78$ | .182 |
| **Ethnicity, n (%)** |  |  |  |  |
| Caucasian | 48 (80%) | 66 (61%) | $\chi^2 = 5.81$ | .016 |
| African | 0 (0%) | 15 (14%) |  |  |
| Asian | 6 (10%) | 2 (2%) |  |  |
| Latino/Hispanic | 3 (5%) | 6 (6%) |  |  |
| Indigenous | 0 (0%) | 6 (6%) |  |  |
| Middle Eastern | 1 (2%) | 5 (5%) |  |  |
| West Indian/Caribbean | 0 (0%) | 3 (3%) |  |  |
| Mixed Race | 2 (3%) | 6 (6%) |  |  |
| **Clinical** |  |  |  |  |
| Age at psychosis onset, years | — | 22.0 ± 4.6 | — | — |
| PANSS Positive (P1+P2+P3) | — | 6.0 ± 3.1 | — | — |
| PANSS Negative (N1+N4+N6) | — | 6.1 ± 3.4 | — | — |
| Antipsychotic dose, DDD | — | 1.21 ± 0.68 | — | — |
| SOFAS | 84.0 ± 4.7 | 58.8 ± 13.8 | t = 16.80 | <.001 |
| TLI Impoverishment | 0.09 ± 0.20 | 0.34 ± 0.39 | U = 1440 | <.001 |
| TLI Disorganization | 0.33 ± 0.40 | 0.48 ± 0.41 | U = 1824 | .010 |
| **Diagnosis** |  |  |  |  |
| Schizophrenia | — | 58 | — | — |
| Schizoaffective | — | 24 | — | — |
| Other Psychosis | — | 22 | — | — |
| Bipolar Disorder | — | 5 | — | — |
| Outpatient/Inpatient status | — | 92/17 | — | — |

**Note.** HC = healthy controls; PT = patients with a psychotic disorder diagnosis. Values are mean ± SD unless otherwise stated. Education is rated on a 1–6 ordinal scale (1 = some primary school; 6 = university degree or higher). SES is rated on a 1–5 ordinal occupational scale (1 = higher/professional occupations; 5 = semi-routine or routine occupations); higher scores indicate lower socioeconomic position. NEET = Not in Education, Employment, or Training. PANSS = Positive and Negative Syndrome Scale; SOFAS = Social and Occupational Functioning Assessment Scale; TLI = Thought and Language Index. Age and SOFAS comparisons used independent-samples t-tests. Sex, NEET, immigrant status, language, cannabis use, and ethnicity were compared using $\chi^2$ tests. Sex is reported as female n (%); male counts were 38 HC and 83 PT. Education, SES, and TLI were compared using Mann–Whitney U tests. Assessment-day antipsychotic defined daily doses (DDD; WHO ATC/DDD index) were available for 96 of 99 patients at baseline. Diagnoses were based on the best-estimate consensus procedure using
DSM-5 criteria. All p-values are two-sided. □ Statistic for Caucasian versus non-Caucasian contrast; individual subgroup cells were too small for formal comparisons.

### 3.1 Group Differences in Conversational Alignment

Register divergence, defined as the within-conversation distance between interviewer and participant word-use profiles, was significantly greater in patient interviews than in control interviews across both timepoints (LMER: β = +0.062, SE = 0.017, *z* = 3.56, *p* = .0004; Cohen’s *d* = +0.48 [0.23, 0.77]; Figure 2). This patient-versus-control difference was greatest during the less-scaffolded early portions of the interview and attenuated in later, more structured sections (see Supplementary Results **SR1** for temporal trajectory; see Supplementary Results **SR2** for decomposition of this signal by individual psycholinguistic dimension; concreteness accounted for 93% of windows reaching significance).

**Figure 2.**
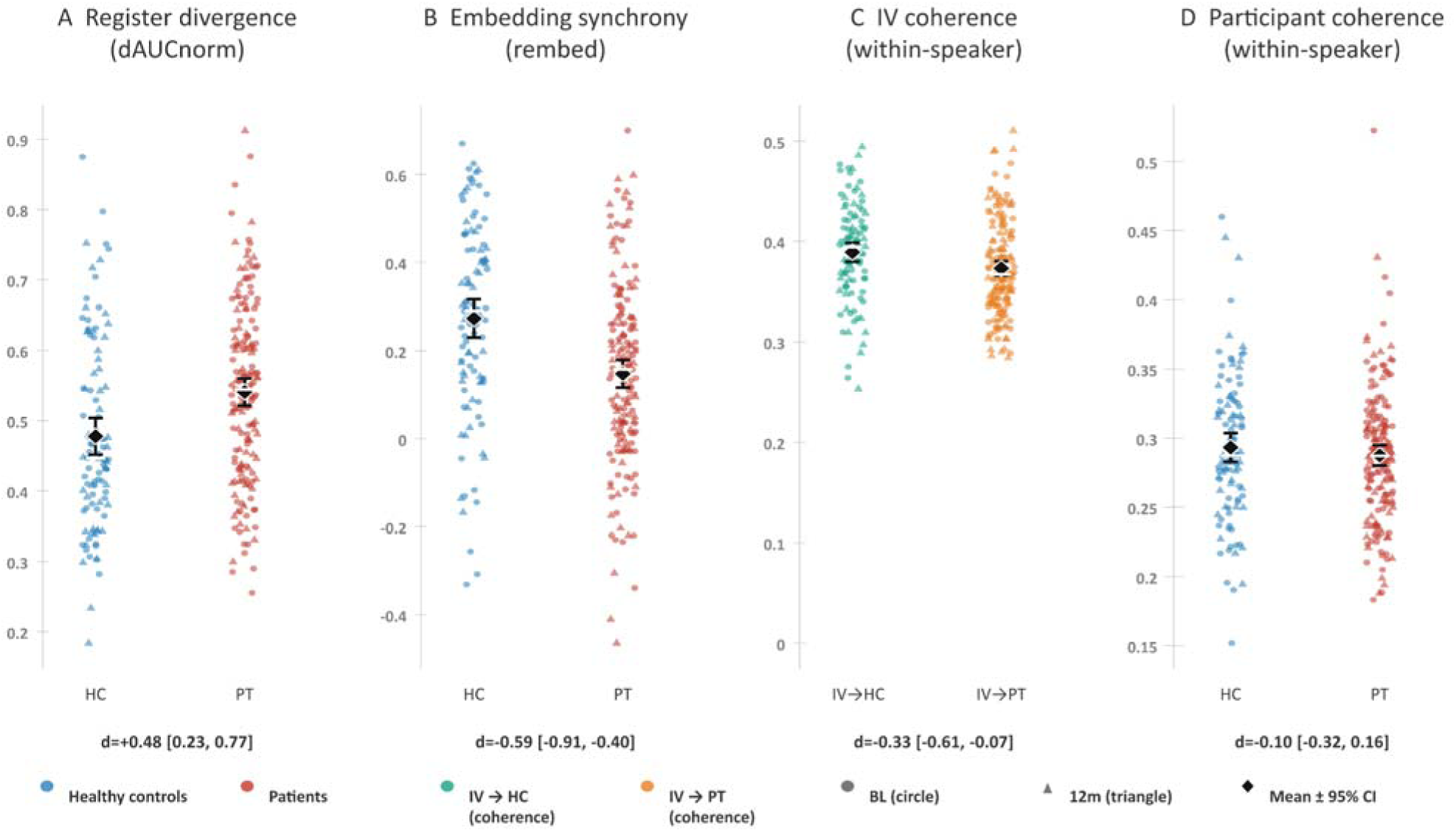
Group differences in conversational alignment metrics at baseline and 12-month follow-up. Each point represents one conversation: circles indicate baseline and triangles indicate 12-month follow-up. Black diamonds and error bars show pooled group means and 95% confidence intervals. Panels A, B and D compare healthy controls (HC, blue) with patients (PT, red); panel C compares interviewer coherence in HC interviews (green) and PT interviews (orange). A: Register divergence (dAUCnorm); higher values indicate greater interviewer–participant divergence in psycholinguistic register. B: Embedding-based synchrony (rembed); higher values indicate stronger coupling between the speakers’ within-speaker continuity streams. C: Interviewer within-speaker coherence. D: Participant within-speaker coherence. The displayed patient–control Cohen’s d estimates and bootstrap 95% confidence intervals correspond to Table 4. There were 102 HC observations and 169 PT observations across both timepoints.

**Table 4.** LMER group comparison results (Baseline + 12m pooled; *n* = 271 observations)

| Metric | HC mean | PT mean | $\beta$ | SE | z | p | d [95% CI] | p×Tp |
| --- | --- | --- | --- | --- | --- | --- | --- | --- |
| Register Divergence | 0.478 | 0.541 | +0.062 | 0.017 | 3.56 | .0004 | **+0.48** [0.23, 0.77] | .914 |
| Embedding-based synchrony | 0.273 | 0.148 | −0.125 | 0.028 | −4.51 | < .001 | **−0.59** [−0.91, −0.40] | .402 |
| Interviewer coherence | 0.389 | 0.373 | −0.015 | 0.006 | −2.42 | .016 | **−0.33** [−0.61, −0.07] | .771 |
| Participant coherence | 0.293 | 0.287 | −0.005 | 0.007 | −0.71 | .482 | −0.10 [−0.32, 0.16] | .146 |

**Note.** $dAUC_{norm}$ = norm-composite register divergence; $r_{embed}$ = sentence-embedding based synchrony; $\beta$ = LMER fixed-effect coefficient (group: patient vs healthy control); SE = standard error; z = Wald statistic; d = LMER-derived Cohen’s d with 95% bootstrap CI (2000 cluster resamples); p×Tp = Group × Timepoint interaction p-value. Metrics in bold reach $p < .05$ .

Participant-level *within-speaker coherence* based on sentence embeddings did not significantly distinguish respondent groups (LMER: *z* = −0.71, *p* = .48, *d* = −0.10). But interviewer’s within-speaker coherence was significantly reduced when the respondents were patients compared to healthy controls (LMER: β = −0.015, SE = 0.006, *z* = −2.42, *p* = .016; *d* = −0.33 [−0.61, −0.07]; Figure 2), highlighting the importance of dyadic dynamics in within-person coherence. *Embedding-based synchrony* with the interviewer was significantly reduced in patient respondents compared to healthy controls (LMER: β = −0.125, SE = 0.028, *z* = −4.51, *p* < .001; *d* = −0.59 [−0.91, −0.40]; Figure 2; Table 4).

These results collectively indicate that patient interviews begin with greater interviewer– participant register divergence and reduced turn-by-turn semantic synchrony, despite preserved participant-level within-speaker coherence in patients; by contrast, interviewer coherence is reduced when conversing with patients compared to healthy participants.

See Supplementary Results **SR6** for turn-structure measures. Patient participants produced more frequent but shorter turns, raising the possibility that differences in conversational structure contributed to the observed alignment effects. To address the longer turns produced by healthy controls, we conducted a sensitivity analysis restricting turns to the first 128 tokens; the primary group differences were unchanged. Also see Supplementary Results **SR7** for reliability-weighted analyses confirming primary findings. This replicated the group differences reported here. See Supplementary Results **SR3** for embedding-based divergence (dAUC_embed_; no HC versus patient difference d = 0.15, p = .278). Complementary speaker-specific analyses in embedding space, including turn-level cross-lagged coupling, are reported in Supplementary Results SR12 (Figure SF10).

### 3.2 Replication across time

Per-timepoint Mann–Whitney tests confirmed the primary outcomes at each assessment. At baseline, register divergence was significantly elevated in patients (r = +0.26, p = .001) and embedding-based synchrony (r_embed_) was significantly reduced (r = −0.33, p < .001). At 12 months, both effects replicated (divergence: r = +0.20, p = .031; r_embed_: r = −0.22, p = .016). No group × timepoint interaction was statistically supported for any metric (all p > .14), providing no evidence that the pooled group differences changed across the 12-month interval. Table 4 summarizes full LMER results.

Interviewer within-speaker coherence was lower in patient than healthy-control interviews at baseline (Mann–Whitney r = −0.23, p = .004), whereas the corresponding 12-month comparison was in the same direction but not statistically supported (r = −0.13, p = .163).

### 3.3 Clinical correlations

Two associations survived BH-FDR correction (Figure 3). Register divergence was positively associated with TLI Impoverishment (partial r = +.17, p = .006, BH-adjusted q = .038); greater impoverishment of speech was associated with greater register divergence from the interviewer, independent of timepoint and participant-specific differences captured by the random intercept. r_embed_ was negatively associated with TLI Disorganization (partial r = −.17, p = .006, q = .038); greater thought disorganization was associated with reduced dyadic synchrony. These associations strengthened with reliability weighting based on the number of turns (Supplementary Results **SR7**). The additional group- and timepoint-adjusted sensitivity analysis showed a positive association between register divergence and TLI Impoverishment, but this did not survive FDR correction. The remaining ten associations did not survive FDR correction. Notable sub-threshold trends included lower r_embed_ and lower SOFAS (partial r = +.14, p = .025) and lower interviewer coherence and higher TLI Impoverishment (partial r = −.10, p = .096; Figure 3). These clinical associations were unchanged after adjustment for assessment-day antipsychotic dose (Supplementary Results SR14), and robustness analyses of the impoverishment association are reported in Supplementary Results SR5.

**Figure 3.**
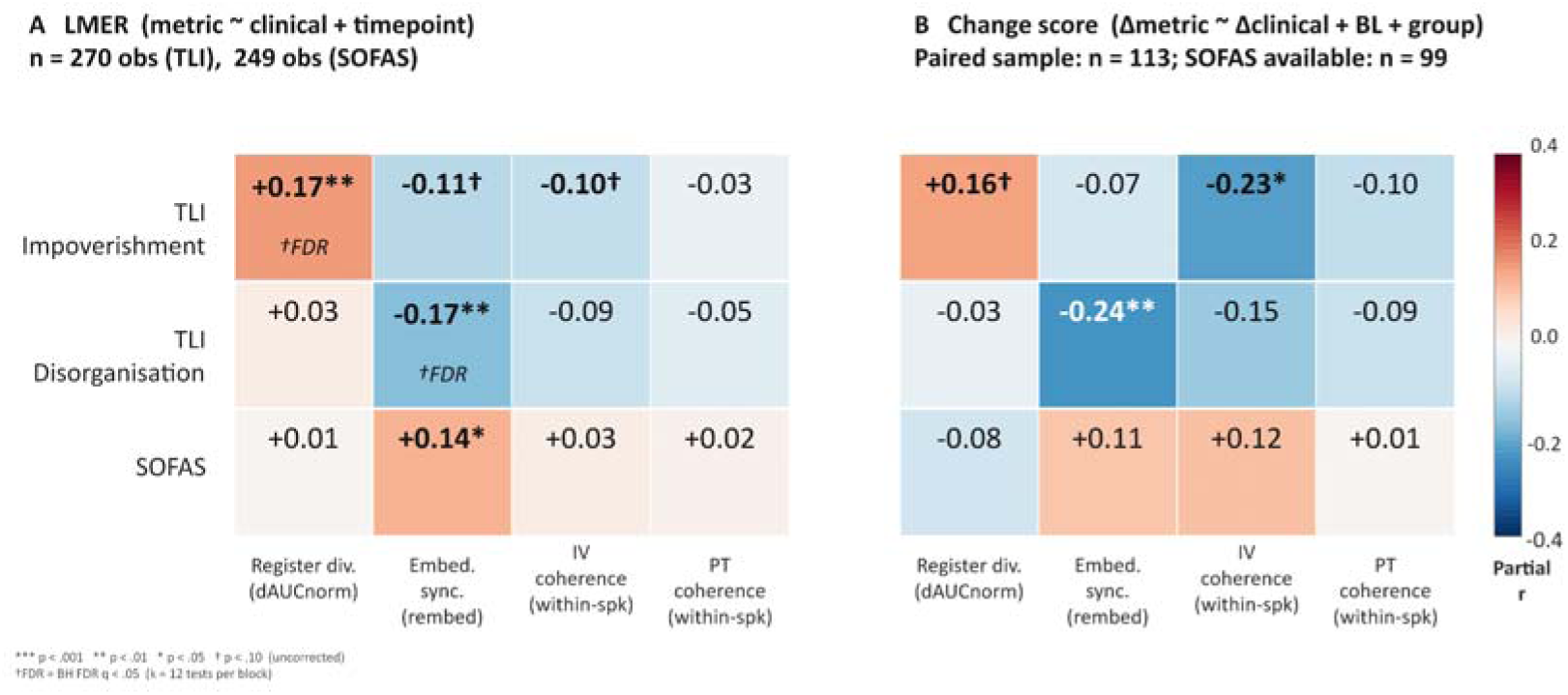
Clinical correlates of conversational alignment. A: Timepoint-adjusted linear mixed-effects associations between four alignment metrics and three clinical variables, with a participant random intercept. B: Associations between within-person changes in alignment and clinical measures, adjusted for baseline values and group. Colour and cell labels show partial r. Stars indicate uncorrected significance (*** p < .001; ** p < .01; * p < .05); † indicates p < .10. The separate †FDR label identifies associations surviving Benjamini–Hochberg correction (q < .05), applied to the 12 tests in each panel. The two FDR-significant associations in panel A are register divergence with TLI Impoverishment and embedding-based synchrony with TLI Disorganization (both q = .038). Neither change-score association highlighted in Results §3.3 survived FDR correction. SOFAS = Social and Occupational Functioning Assessment Scale; TLI = Thought and Language Index; IV = interviewer; PT = participant.

Change-score analyses tested alignment–clinical covariation in 113 paired participants. Embedding-based synchrony (Δ*r*_embed_) was negatively associated with ΔTLI Disorganization (partial *r* = −0.24, *p* = .010) and Δinterviewer coherence was negatively associated with ΔTLI Impoverishment (partial *r* = −0.23, *p* = .018). Neither association survived BH-FDR correction (both *q* > .06). These change-score associations should be considered exploratory trends directionally consistent with the primary cross-sectional analysis. Supplementary Results **SR5** presents severity-bin visualizations and the High-Impoverishment robustness analysis.

When individual DSM-5 diagnoses were considered, patients with schizophrenia showed significantly greater register divergence than other psychotic disorder diagnoses (Supplementary Results **SR9**); hospitalization status did not substantially moderate alignment profiles (Supplementary Results **SR10**).

## 4. Discussion

Semi-structured interviews involving patients with psychotic disorders showed misattunement across three computationally distinct linguistic dimensions. First, patient interviews showed greater interviewer–participant register divergence, driven predominantly by the concreteness of patients’ lexical choices. This pattern is consistent with a more concrete and restricted word-use profile associated with impoverished thinking that diverged from the interviewer’s more varied register. Second, moment-by-moment coupling of semantic trajectories was reduced, with embedding-based synchrony tracking TLI Disorganization rather than Impoverishment. Third, interviewer within-speaker coherence was reduced during patient interviews relative to healthy-control interviews, suggesting that linguistic phenomena of psychosis were not confined to patient speech alone but extended to include the conversational dyad.

Reduced interviewer coherence may reflect interactional demands associated with impoverished speech, including increased prompting or reformulation in response to reduced patient elaboration. It may also arise from adaptive accommodation or conversational repair ^(65,66)^, which the present analyses do not distinguish. Nonetheless, the selective alteration of interviewer coherence during patient interviews demonstrates that the communicative difference is organized at the dyadic level, regardless of whether the interviewer’s response is adaptive or maladaptive. Future studies should incorporate instructions for greater reciprocal exchange with substantive contributions from both speakers to distinguish the direction of conversational adaptation.

Reduced conversational synchrony indicates weaker turn-by-turn coupling of semantic trajectories, reflecting impaired coordination of meaning across successive contributions. Cooperative conversation requires each speaker to use the other’s prior contribution to constrain what comes next, while suppressing irrelevant or distracting associations ^(38,67)^. When this coordination is reduced, the exchange may lose continuity even when individual contributions remain locally coherent ^(36,68–70)^. This extends the interactive alignment framework ^(37)^ to interviews in psychosis, moving from structural matching paradigms ^(47,48)^ to broader semantic embedding levels in clinical exchanges.

The findings have several methodological and clinical implications. The within-interview profile of register divergence indicates that group differences were largest during the more open-ended, less scaffolded early portions of the interview and attenuated during later structured sections. This finding suggests that external task structure may provide shared referential anchors that reduce observable register divergence. Counterbalanced task designs are needed to distinguish the effects of task structure from conversational familiarity. One implication for conversational assessments is that the less scaffolded discourse in psychosis appears more sensitive to communicative misalignment, whereas highly structured tasks may underestimate (‘mask’) interactional disruption.

Alignment metrics showed longitudinal persistence, with patient–control differences remaining detectable at 12-month follow-up and stronger register divergence associated with schizophrenia diagnosis. However, these group-level findings do not establish robust within-person state sensitivity or individual-level prediction of clinical course, which require larger repeated-measures samples with wider ranges of clinical severity.

Several limitations should be acknowledged. Patients produced shorter, more numerous turns, and this turn structure was associated with impoverished speech; however, sensitivity analyses suggested that the main findings were not driven by differences in turn structure. Transcription was semi-automated using Batchalign with manual verification, so the current pipeline cannot yet be treated as a transcript-to-results workflow for clinical deployment. Finally, the sample was predominantly young, English-speaking, predominantly recent-onset, and assessed at a single Canadian site using a specific semi-structured protocol. Thus, generalizability to chronic schizophrenia, other interview formats, languages, and neuropsychiatric conditions such as autism or frontotemporal dementia remains to be established. Audio-derived prosodic features were not included; integrating acoustic and lexico-semantic signals is therefore an important next step toward a fuller multiscale account of interactional attunement. In addition, most interviews were conducted by a single trained interviewer; the interviewer-coherence findings therefore describe one interviewer’s conversational behaviour and require replication with more interviewers.

These transcript-derived alignment metrics provide a bridge toward an account of interpersonal attunement. Combining these with dual-brain EEG hyperscanning ^(32,71,72)^ and computational movement tracking would enable simultaneous quantification across lexical, acoustic, neural, and embodied scales within a single dyadic exchange. Although the *praecox feeling* was not operationalized here, future studies combining these measures with contemporaneous interviewer ratings could test whether variation in interviewer speech corresponds to subjective evaluations of the interaction. Because they are derived from plain-text clinical interview transcripts without specialized equipment, these metrics offer a scalable, low-burden approach to studying communicative function, with potential applications for predicting social functioning and digital phenotyping. With prospective cross-linguistic, longitudinal, and cross-context validation across psychotherapy, routine clinical encounters, and unconstrained conversation, they may provide candidate computational readouts of interpersonal communication in psychosis.

## Data availability statement

The data used in the study are available through the DISCOURSE in Psychosis consortium’s Psychosis Talkbank https://psychosis.talkbank.org/. Requests to access the datasets should follow the instructions on the website.

## Acknowledgments

Special thanks to Mike Mackinley and Sabrina Ford for data curation and transcription support.

## Author Contributions

**Emmanuel Olarewaju:** Conceptualization; Methodology; Investigation; Formal analysis; Validation; Interpretation; Writing – original draft; Writing – review & editing; Visualization; Project administration. **Alban Voppel:** Curation; Writing – review & editing. **Fiona Meister:** Conceptualization; Writing – review & editing. **Chaimaa El Mouslih:** Curation; Writing – review. **Paulina Dzialoszynski:** Curation. **Lena Palaniyappan**: Supervision; Funding acquisition; Resources; Conceptualization; Methodology; Investigation; Formal analysis; Validation; Interpretation; Writing – original draft; Writing – review & editing; Visualization.

## Funding

Lena Palaniyappan’s research is supported by Monique H. Bourgeois Chair in Developmental Disorders and the Graham Boeckh Foundation. He receives a salary award from the Fonds de recherche du Québec-Santé (FRQS: 366934) and supported by the FRQS through a Research Centre Grant to Douglas Research Centre (https://doi.org/10.69777/5230). Funding was also provided to Emmanuel Olarewaju by Healthy Brain, Healthy Lives (HBHL) Fellowship and Graduate Excellence Recruitment award from McGill University. Alban Voppel is supported by NARSAD Young Investigator Grant 32574 from the Brain & Behaviour Research Foundation. Chaimaa El Mouslih is supported by the Fonds de recherche du Québec-Société et Culture (grant # 370599).

This work is supported by the FRQS Partenariat Innovation-Québec-Janssen (PIQ-J) initiative (#338282 awarded to LP: https://doi.org/10.69777/338282) and FRQS Alliance en santé mentale grant (#348797 awarded to Douglas Research Center); Canadian Institutes of Health Research (CIHR) - Strategy for Patient Oriented Research Priority Announcement (SPOR; Grant number PJK192157) and Project Grant (Grant number PJT195903); Wellcome Trust Discretionary Grant (226168/Z/22/Z to Dr. Iris Sommer and LP) and Mental Health Award for the DIALOG consortium (314138/Z/24/Z to LP)

## Conflicts of interest

The authors report no conflict of interest

## Declaration of generative AI use

During the preparation of this manuscript, the authors used OpenAI Codex (v5.5) and Anthropic Claude Opus (v4.7), accessed in 2026, to assist with code development, debugging, visualization, data-analysis verification, and manuscript editing. The authors reviewed, verified, and edited all AI-assisted outputs and take full responsibility for the accuracy and integrity of the final manuscript, analyses, and conclusions.

## Software and computational environment

All analyses were performed in Python 3.10.12. Tabular data were read from Microsoft Excel and CSV formats using pandas 2.3.3, openpyxl 3.1.5, and numpy 2.2.6. Sentence embeddings were generated using the sentence-transformers library 2.7.0. Linear mixed-effects models were estimated using statsmodels 0.14.6, and additional regression-based analyses were performed using scipy 1.15.3 and scikit-learn 1.7.2. Figures were produced using Matplotlib 3.10.8.

## Ethics statement

The study was reviewed and approved by the Western University Health Sciences Research Ethics Board. The patients/healthy controls provided their written informed consent to participate in the study.

## Supplementary Materials

### SM1 Relevance of register-level psycholinguistic norms to the study of schizophrenia

For the register level, psycholinguistic norms of word meaning provide a principled basis for operationalization, with dimensions selected on the basis of established mechanisms of language disturbance in schizophrenia. Patients characteristically show reduced type–token ratio and lexical diversity ^(73–80)^, i.e., vocabulary restricted to a narrower, more repetitive range that concentrates word choice in the high-frequency, semantically non-specific core of the lexicon. This pattern is captured by two complementary norm dimensions. A. Semantic neighbourhood density ^(59)^ indexes how many words share a similar distributional position with a target in corpus co-occurrence space: high-density words are frequent, contextually generic terms that appear across many discourse settings, and a speaker restricted to this high-frequency core diverges systematically from an interviewer who deploys contextually varied, specific vocabulary. B. Semantic diversity ^(58)^ quantifies how broadly distributed a word’s contexts of use are across the corpus; a failure to exploit discourse context to constrain lexical selection (^81^; also see ^82–84^) produces word choices that appear unrestricted to any particular topic or situation, yielding a register that feels ambiguous and topically underdetermined to the listener.

Patients with schizophrenia exhibit difficulties in abstract thinking and resort to concrete interpretations of language ^(85,86)^. They produce specific, perceptually grounded referents infrequently, and instead rely on vague, context-general function words ^(74,87,88)^. Concrete concepts carry both verbal and sensorimotor representations, making them more reliably accessible under contextual demand. Norms of concreteness, along with visual and auditory perceptual strength in the sensorimotor domain, capture the abstractness level at which the conversational register is anchored in a speaker.

Taken together, the five dimensions based on lexical choice (neighbourhood density and diversity) and concreteness register (abstract and visual/auditory perceptual grounding) define a composite semantic register that has been shown by Sacks and colleagues to be sensitive to dyadic conversational alignment and remain tractable, unlike LLM-based high dimensional vectors (see ConversationAlign ^56^).

### SM2 Data availability

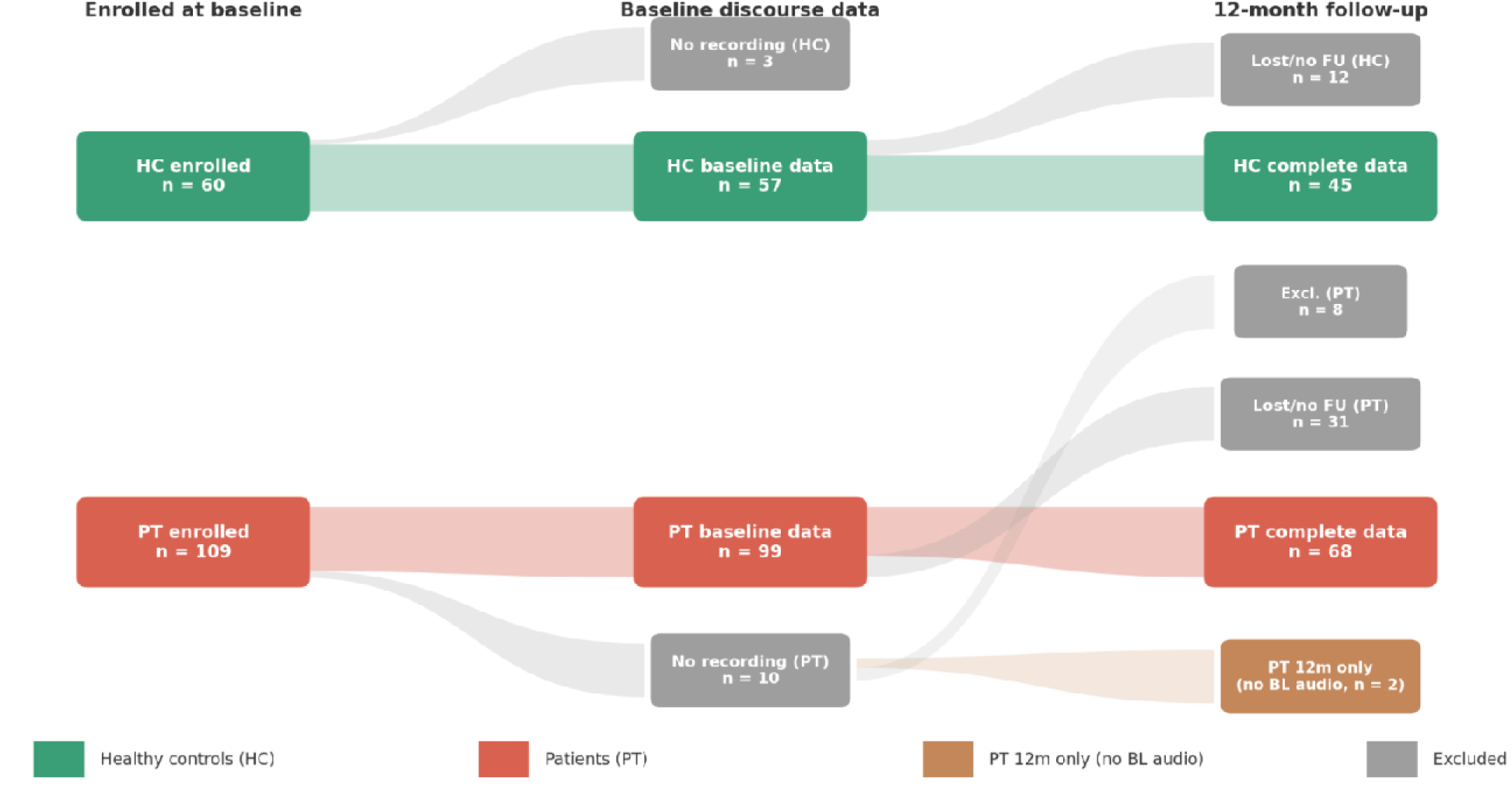

***Note.*** Flow diagram depicting participant enrollment, baseline discourse data availability, and 12-month follow-up outcomes for healthy controls (HC) and patients (PT). Attrition reflects loss to follow-up and recordings unavailable at either time point. Counts are shown for each stage, with complete cases defined by availability of usable discourse data at both baseline and follow-up.

### SM3 Interview protocol

The DISCOURSE protocol is a semi-structured interview that uses a fixed sequence of discourse contexts but does not prescribe conversational content or turn-by-turn exchanges. It includes a free conversational speech section, in which participants introduce themselves and discuss daily life, work, family, and personal interests; a personal narrative section, in which they recount a significant life event in their own words; a picture-description task, requiring detailed descriptions of images; and a storyboard-narration task, requiring study of an illustrated sequence and retelling of its story in the participant’s own words. The protocol also includes health narrative, dream report, and reading-and-recall sections. Section order, task instructions, and 14 initiating interviewer prompts/questions were standardised.

Apart from the prescribed reading-aloud component, exchanges were unscripted and contingent on participants’ contributions. Interviewers selected follow-up topics and questions in response to those contributions, and could rephrase questions or provide non-directive prompts, while otherwise minimising verbal intervention. Thus, the protocol standardised elicitation while allowing unscripted, responsive exchanges within each discourse context. Each interview was administered in a single session by a trained research interviewer. Follow-up prompts (e.g., “Can you tell me more?” or “Anything else?”) were used only when a participant paused for more than 10 seconds or did not initiate speech within 30 seconds; they were otherwise withheld.

**Table ST1.** DISCOURSE interview task structure.

| # | Task | Description |
| --- | --- | --- |
| 1 | Unstructured<br>Conversation | Open-ended dialogue about daily life, occupation, and interests. No external structure; register selection is spontaneous. |
| 2 | Personal<br>Narrative | Participants recall and describe a significant recent or past life event. Episodic memory and autobiographical discourse. |
| 3 | Health Narrative | Participants discuss their mental and physical health history. Quasi-clinical register; both parties converge on a health-discourse domain. |
| 4 | Picture Description (3 TAT) | Three Thematic Apperception Test pictures presented sequentially (1 min per picture). Participant describes what they see and infers what is happening. Demands social-inferential language and theory-of-mind reasoning. |
| 5 | Storyboard (Lighthouse) | Six-panel wordless storyboard. Participant constructs and narrates the depicted event sequence with a shared visual referent. |
| 6 | Dream Reports | Participants describe a recent or recurring dream, requiring hedged subjective framing of non-real events. |
| 7 | Reading and Recall | Participants read a one-page passage aloud (Pitcher and Crow story), then retell it in their own words. Reading provides articulatory data; recall assesses memory-driven discourse planning. |

### SM4 Transcription process

#### Transcript format

Transcriptions were generated with *Batchalign*, a Python program that can create CHAT transcripts from recorded audio files. The generated transcripts were then manually verified by two researchers (MM, CEM) with the original audio recordings using the CLAN program. Original audio files and the verified transcripts were then uploaded to PsychosisBank servers. For the current analysis, these transcripts were downloaded and stored as plain-text files (.txt), and speaker diarization was cross-checked using whisperX diarization with manual checking, each line of a transcript carries a speaker prefix followed by the text of a single spoken utterance:

Interviewer: [utterance text]

Participant: [utterance text]

An utterance, as used in the transcripts, corresponds to a natural breath-group or pause unit as judged by *Batchalign* and manual checks. Consecutive lines from the same speaker that occur before the other speaker begins are each written as separate lines. No timing information, prosodic markings, or non-verbal events appear in the files used for the present analysis (timestamped versions of the transcripts are available but not used here). All interviews returning a valid plain-text file were included, provided the automatic parsing step identified at least three interviewer turns and at least three participant turns (see below for the definition of turns). This minimum was chosen to ensure that turn-by-turn trajectory metrics would be numerically stable. Participants with fewer than three turns per speaker were planned to be excluded from all analyses, but no such cases were found.

#### Turn Segmentation and Merging

##### Definition of a turn

The turn, not the individual sentence, is the fundamental unit of conversational exchange. Turns typically consist of several utterances as natural conversations are rarely in concrete sentences and may span several words. For our study, a turn is defined as an uninterrupted stretch of speech by one speaker before the other speaker begins, regardless of how many utterance lines it spans in the transcript. Operationally, consecutive lines sharing the same speaker prefix are merged by concatenating their text with a single space. A new turn begins whenever the speaker prefix changes.

For example, these three consecutive transcript lines:

Participant: I am a social worker.

Participant: I recently started my career.

Participant: It has been a big change.

are merged into a single participant turn: *“I am a social worker. I recently started my career. It has been a big change.”*. This entire passage is treated as one unit for all downstream computations. For all transcripts, at this stage of term definition, a manual check was carried out (LP) to ensure that no inadvertent labelling of a participant’s utterance as an interviewer’s utterance has occurred during the process. Treating each line as an independent turn would artificially inflate turn counts and fragment semantically coherent passages. Merging lines from the same speaker restores the natural turn structure of the conversation. Of note, a turn transition occurs at every change of speaker, including single-word back-channel responses (such as “Yeah.” or “Okay.”) by the interviewer. Back-channels were not excluded because they are part of the natural rhythmic structure of conversation. The metrics are designed to be computed over the paired sequence of interviewer and participant turns, and removing back-channels would distort this structure. This also allowed us to assess how long (in terms of number of words or tokens) a speaker holds the conversational floor across multiple utterances without interruption, and the number of turns taken by each person in the conversational dyad.

#### Text Preprocessing

The register divergence metric (dAUC_norm_) is based on psycholinguistic word norms. Only this metric requires text preprocessing; the sentence-embedding metrics (see below) use the raw, unprocessed turn text. The psycholinguistic norm database contains entries for English words in lowercase alphabetic form, including words with internal apostrophes such as “can’t” and “it’s”. To match this, each turn was converted to lowercase and tokenised retaining only sequences of lowercase letters and apostrophes, discarding numbers, punctuation, and symbols. The chosen pattern captured all contracted forms while excluding tokens that cannot be looked up (numerals, filler syllables transcribed as non-alphabetic strings). Single-character tokens were discarded in the filtering stage. Each token was lemmatised using NLTK’s WordNet Lemmatizer, which maps inflected word forms to their dictionary base form (e.g., “talking” → “talk”). Lemmatisation is applied before and after stopword filtering to ensure that both inflected and base forms of stopwords are removed using a combined stopword list. The list merges standard English stopwords (function words, pronouns, prepositions, auxiliary verbs) with a custom set of common conversational fillers (“yeah”, “okay”, “um”, “uh”, etc.). Dual-pass filtering was applied i.e., once before lemmatisation (to the raw token) and once after lemmatisation (to the lemmatised form). The output of this pipeline was a set of content words (primarily nouns, verbs, adjectives, and adverbs) for each turn to be looked up in the psycholinguistic norm database. Turns that produce zero content words (e.g., a turn consisting entirely of filler words e.g., “Yeah, okay”) were assigned a missing value for the norm composite at that position.

### SM5 Alignment Metrics

#### Summary of conversational alignment metrics and units of analysis

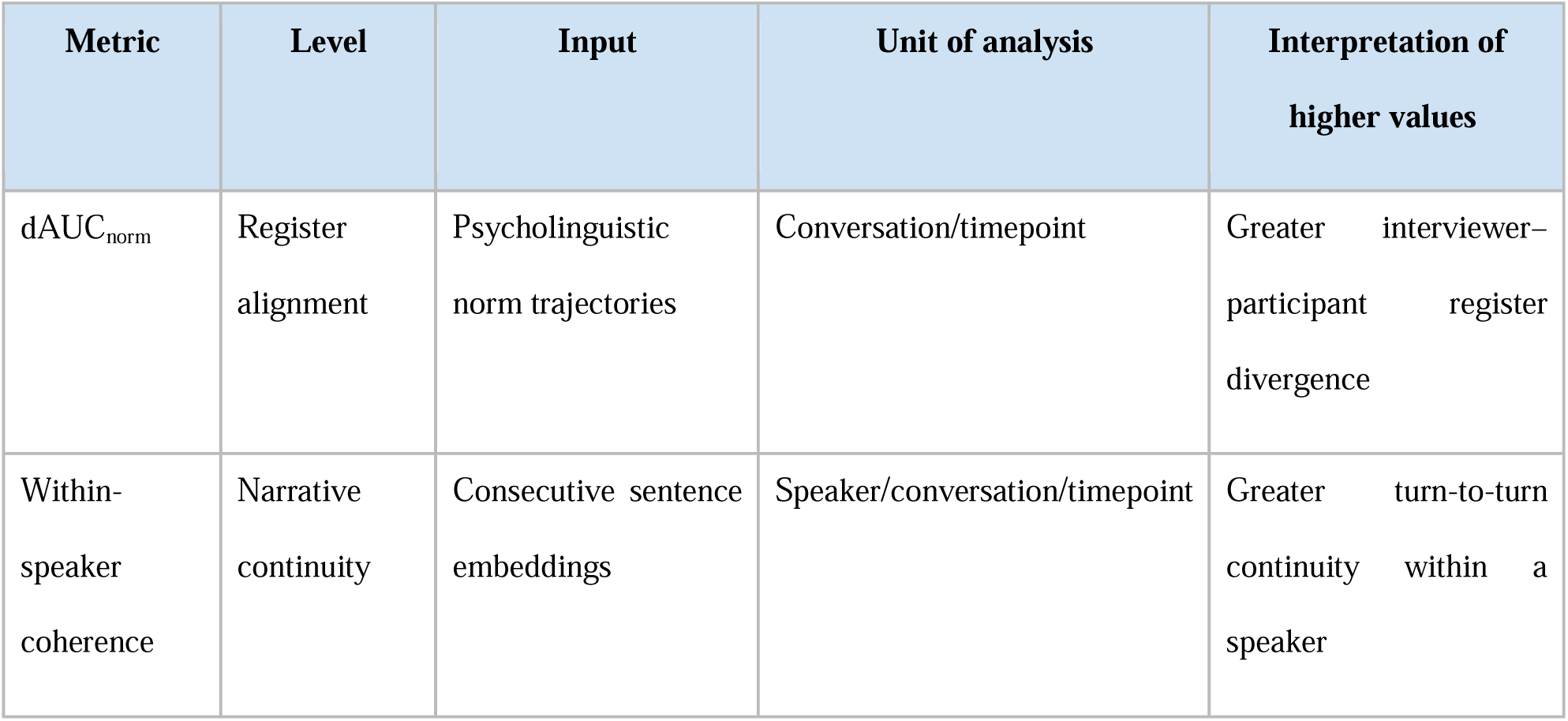
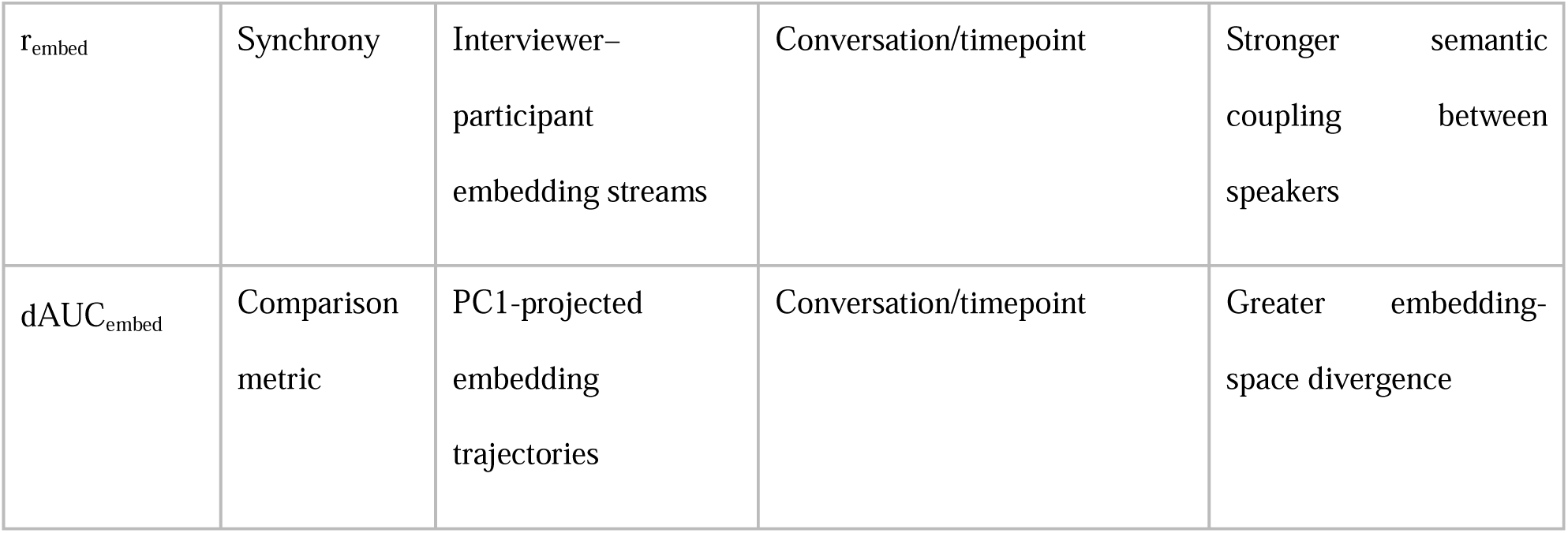

#### Verbatim examples of register alignment (dAUC_norm_)

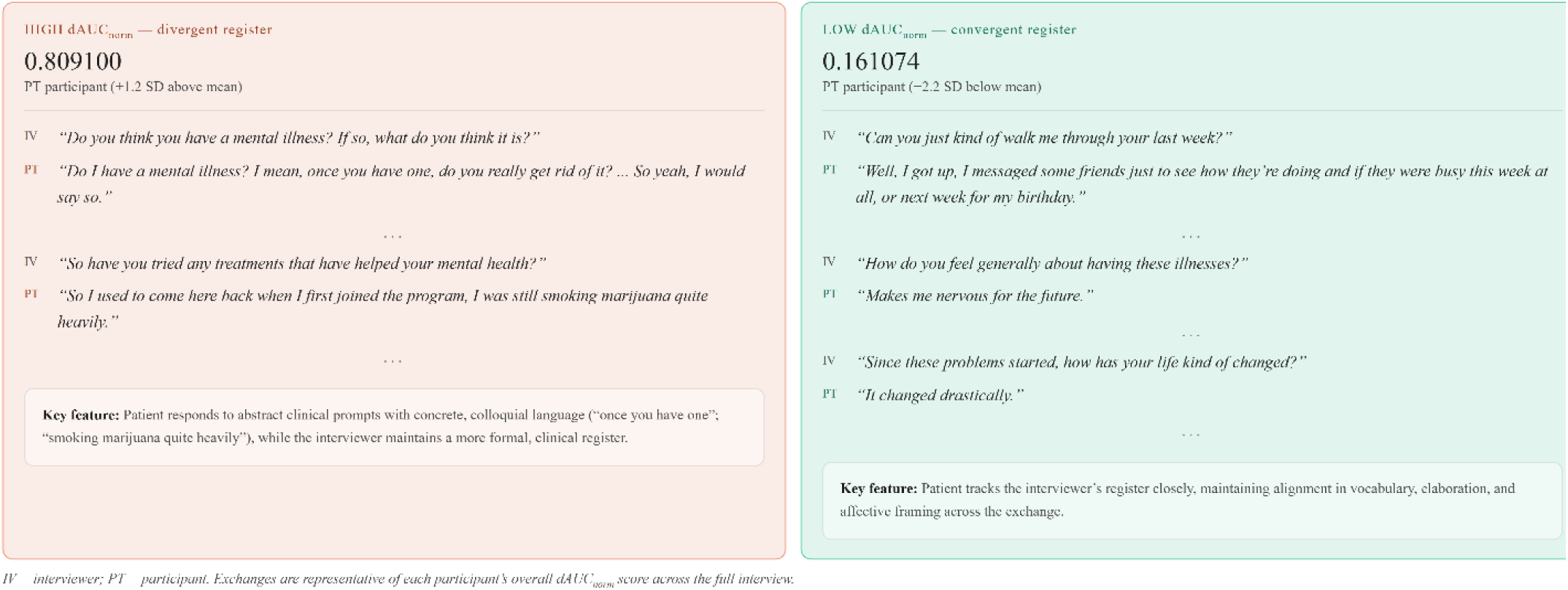

***Note.*** IV = interviewer; PT = participant. Register divergence (dAUC_norm_) indexes accumulated turn-by-turn distance between interviewer and participant lexical-semantic register profiles across the full interview. Higher values indicate greater register dissimilarity, whereas lower values indicate greater register similarity. The displayed exchanges are illustrative and are not themselves the basis of the conversation-level score. Register similarity does not establish directional convergence, accommodation, or causal influence.

#### Verbatim examples of embedding-based synchrony

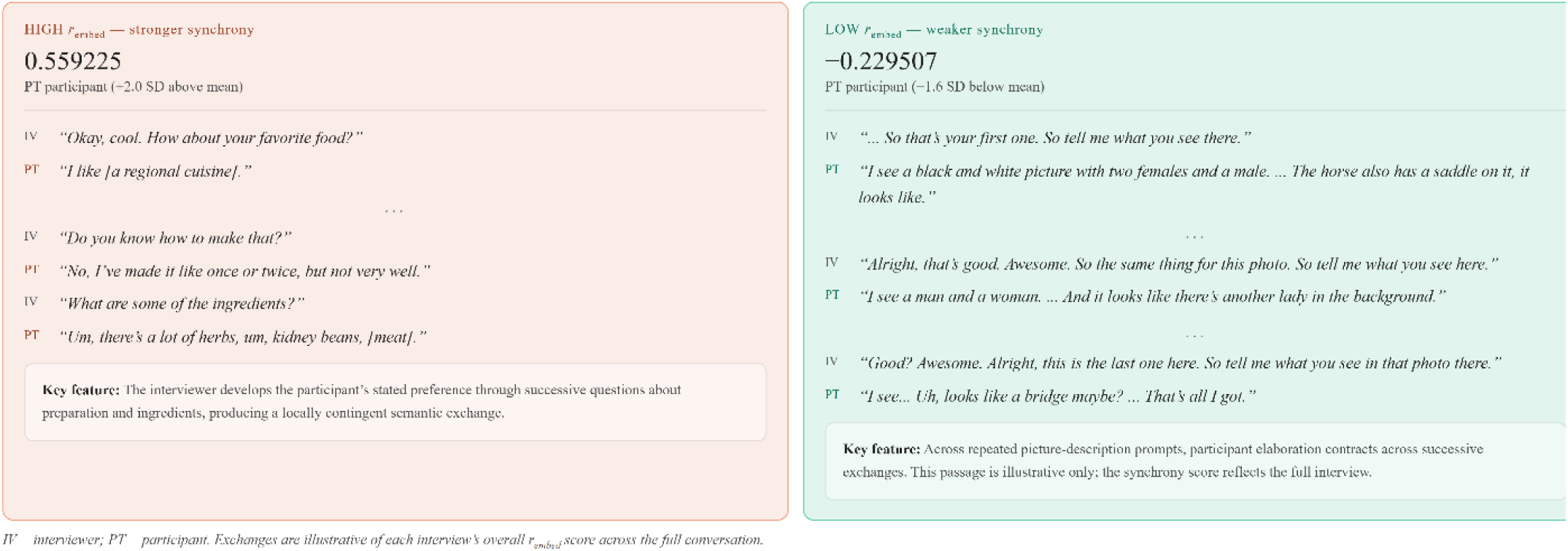

***Note.*** IV = interviewer; PT = participant. Embedding-based synchrony (r_embed_) indexes positive coupling between the speakers’ sentence-level semantic-continuity streams across the full interview. Higher values indicate stronger temporal coordination of semantic development but do not require lexical similarity in every adjacent exchange. The displayed exchanges are illustrative and are not themselves the basis of the conversation-level score.

#### Verbatim examples of within-speaker coherence

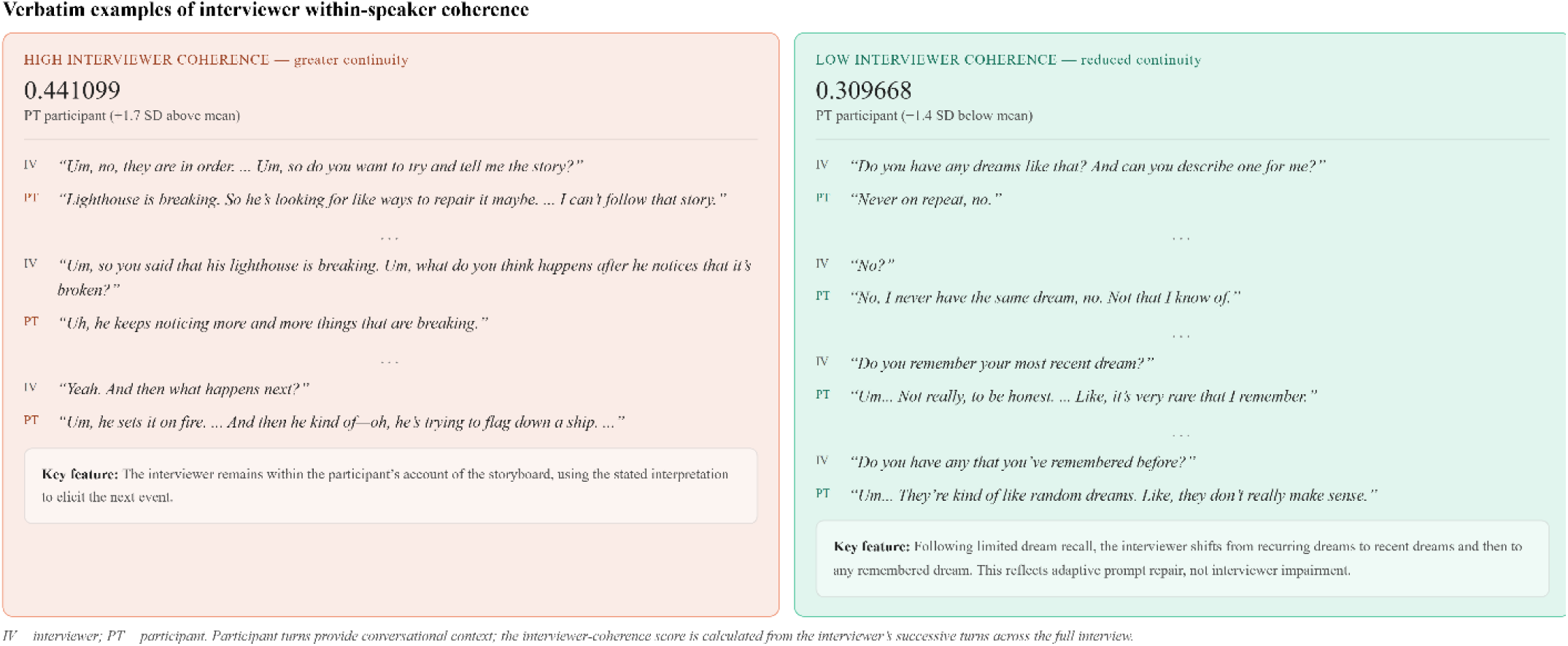

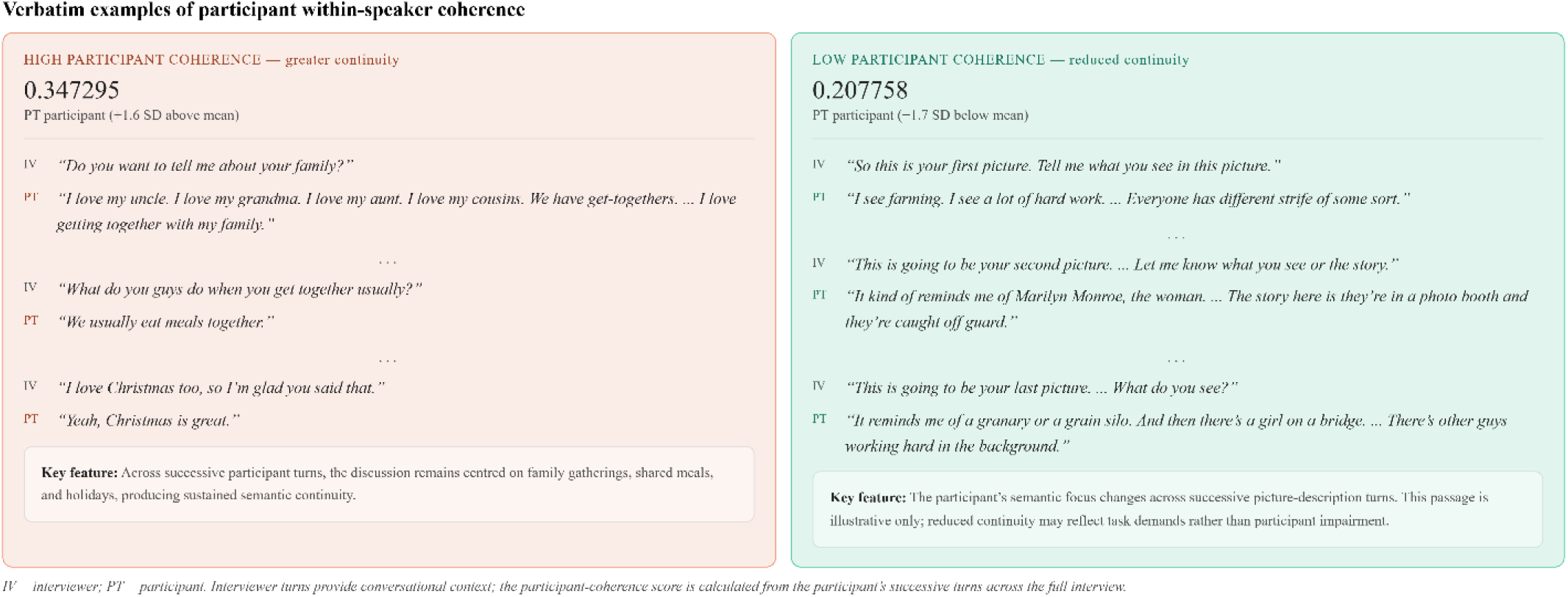

***Note.*** IV = interviewer; PT = participant. The upper panel presents interviewer within-speaker coherence, calculated from semantic continuity across the interviewer’s successive turns; participant turns are shown only to preserve conversational context. The lower panel presents participant within-speaker coherence, calculated from semantic continuity across the participant’s successive turns; interviewer turns provide context. Higher values indicate greater semantic continuity within the focal speaker’s turn sequence. Lower values may reflect task transitions, repair, prompting, response demands, or other conversational processes. The displayed exchanges are illustrative and are not themselves the basis of the full-interview scores.

##### Norm database

The database contains psycholinguistic and sensorimotor norm values for approximately 156,000 English words. Five semantic dimensions are used for this analysis: Auditory experience (sem_auditory_rescale): *Extent to which the word evokes auditory imagery*. Concreteness (sem_cnc_b24_rescale): *Degree to which the word refers to a tangible, physical entity*. Semantic diversity (sem_diversity_rescale): *Breadth of contexts in which the word is used across a large text corpus*. Number of associates (sem_neighbors_rescale): *Number of semantic neighbours (free-association strength)*. Visual experience (sem_visual_rescale): *Extent to which the word evokes visual imagery*.

For each turn, the set of content words is looked up in the database. For each of the five dimensions, the mean value across all words that have a non-missing entry for that dimension is computed. These five dimension means are then averaged to produce a single per-turn composite value (aggregated per-turn norm composite). To avoid unstable averaging from sparse coverage, a composite was only computed if at least three of the five dimensions are represented. This composite represents the average psycholinguistic register value of the turn.

##### Sentence-Embedding Metrics

Three of the four primary metrics use dense vector representations of turns produced by a multilingual sentence encoder. These metrics [sentence-embedding synchrony (*r*_embed_), interviewer within-speaker coherence (iv-cont-embed), and participant within-speaker coherence (pt-cont-embed)] operate on raw, unprocessed turn text, with no stopword removal or lemmatisation. The model used is paraphrase-multilingual-MiniLM-L12-v2 from the SentenceTransformers library ^(61)^. It is a 12-layer distilled transformer with a 384-dimensional output space. The model was selected because it was pre-trained for semantic similarity tasks across multiple languages, making it appropriate for future studies in other languages in the Psychosis Talkbank. Each turn is encoded as a single 384-dimensional dense vector by mean-pooling the token representations from the final transformer layer. Note that each *merged turn* (as defined earlier) - not a single sentence or utterance - is passed to the model as a single input sequence. Metrics comparing interviewer turns to participant turns (such as synchrony and register divergence) require a single vector per turn, not one per sentence. Encoding at sentence level and then averaging would discard the sequential structure within turns and would produce vectors of inconsistent reliability (a one-sentence turn and a ten-sentence turn would receive equal treatment). Using the full turn as input allows the model to integrate semantic content across all utterances that make a turn when producing the summary vector.

Transformer models internally represent input as a fixed-length sequence of sub-word tokens. Turns longer than the model’s maximum sequence length are truncated: the portion beyond the limit is discarded before encoding. Sub-word tokenisation typically produces more tokens than words (on average about 1.2–1.5 sub-word tokens per word in English), so a 512-token limit (rather than the default of 128) that accommodates roughly 350–420 words, large enough to accommodate most turns in the Discourse speech tasks protocol, was used. Of note, healthy controls produced substantially longer turns than patients. Approximately 13% of healthy-control turns exceeded the 128-token limit at baseline, compared with approximately 6% for patients. This differential truncation would artificially compress the tail of healthy-control embedding distributions, attenuating the group difference in metrics that depend on full turn content. The use of 512-tokens ensured <0.1% truncation for both groups, confirming that essentially the full content of every turn was encoded. We undertook a 128-token sensitivity analysis (Table ST7) to assess whether the primary inferential conclusions were preserved under the shorter limit.

The quantitative change is that sentence-embedding synchrony (*r*_embed_) yields a larger group effect with 512-token encoding (LMER Cohen’s *d* = −0.59 versus −0.50 at 128 tokens), consistent with the prediction from the truncation analysis.

##### Metric Computation

All metrics are computed per conversation (one value per participant per timepoint) from the turn sequences produced as per the steps above.

##### Register Divergence (dAUC_norm_)

The interview is treated as a sequence of paired exchanges: the first interviewer turn is paired with the first participant turn, and so on up to *n* = min(number of interviewer turns, number of participant turns) pairs. At each exchange position *t*, the absolute difference between the interviewer’s norm composite and the participant’s norm composite is computed. The area under this turn-by-turn difference curve is estimated by the trapezoidal rule and divided by *n* − 1 to normalise for conversation length:

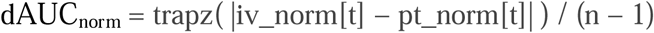

Higher values indicate greater divergence in psycholinguistic register. Positions where either speaker’s composite is missing are excluded from the integration.

##### Embedding-based Synchrony (r_embed_)

The within-speaker continuity stream for each speaker is computed: for the interviewer, this is the cosine similarity between each consecutive pair of interviewer turn vectors. For *n* interviewer turns, this yields a continuity series of length *n* − 1. An equivalent series is computed for the participant. The two series are then correlated (Pearson *r*) up to the length of the shorter series:

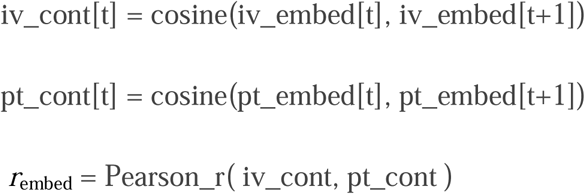

Higher values indicate that the interviewer’s sequential semantic self-consistency co-varies positively with the participant’s, reflecting rhythmic synchrony in how the two speakers move through semantic space.

##### Within-Speaker Coherence (iv-cont-embed and pt-cont-embed)

For each speaker, the mean of the continuity stream (as defined above) is taken as the within-speaker coherence score:

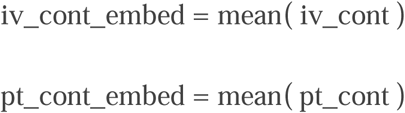

Higher values indicate that a speaker’s consecutive turns are more semantically similar on average. In other words, the speaker tends to remain on topic from one turn to the next. The interviewer coherence metric (iv-cont-embed) captures how consistently the interviewer maintains a conversational thread, and is interpreted as measuring the coherence demands of speaking with a given partner.

##### Embedding-Based Divergence (dAUC_embed_)

For dAUCembed, PCA was used only as a dimensional-reduction step for a supplementary embedding-space comparison metric. Within each conversation, interviewer and participant turn embeddings were stacked before PCA, and only PC1 was retained by design to yield a single scalar trajectory parallel to the norm-based divergence trajectory. Each turn was projected onto PC1, and dAUC_embed_ was computed as the normalized trapezoidal integral of the absolute interviewer-participant difference across paired turns. Additional components were not included in the primary dAUC_embed_ computation, and the main register-divergence finding does not depend on PCA because it was derived from the five-dimensional psycholinguistic norm composite.

### SM6 Statistical Analysis

#### Linear mixed-effects models (LMER)

All group comparisons and clinical associations were tested using linear mixed-effects models implemented in the Python *statsmodels* library, with the model formula:

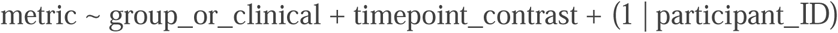

where the participant-level random intercept accounts for the two-observation repeated-measures structure (baseline and 12-month assessments nested within participants). Timepoint was entered as a deviation-coded contrast (baseline = −0.5, 12-month = +0.5).

#### Multiple comparison correction

For clinical associations, the set of 12 tests in each analysis family (4 alignment metrics × 3 clinical variables) was corrected using the Benjamini-Hochberg false discovery rate procedure at a nominal rate of 5%. The LMER and change-score analyses were treated as independent testing families and corrected separately.

#### Cohen’s d

LMER-derived effect sizes were computed as the ratio of the group fixed-effect coefficient to the square root of the sum of the residual variance and half the random-effect variance:

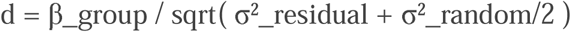

95% confidence intervals on *d* were estimated from 2,000 cluster bootstrap resamples drawn at the participant level (i.e., all observations from a given participant were included or excluded together).

**Table ST2:**
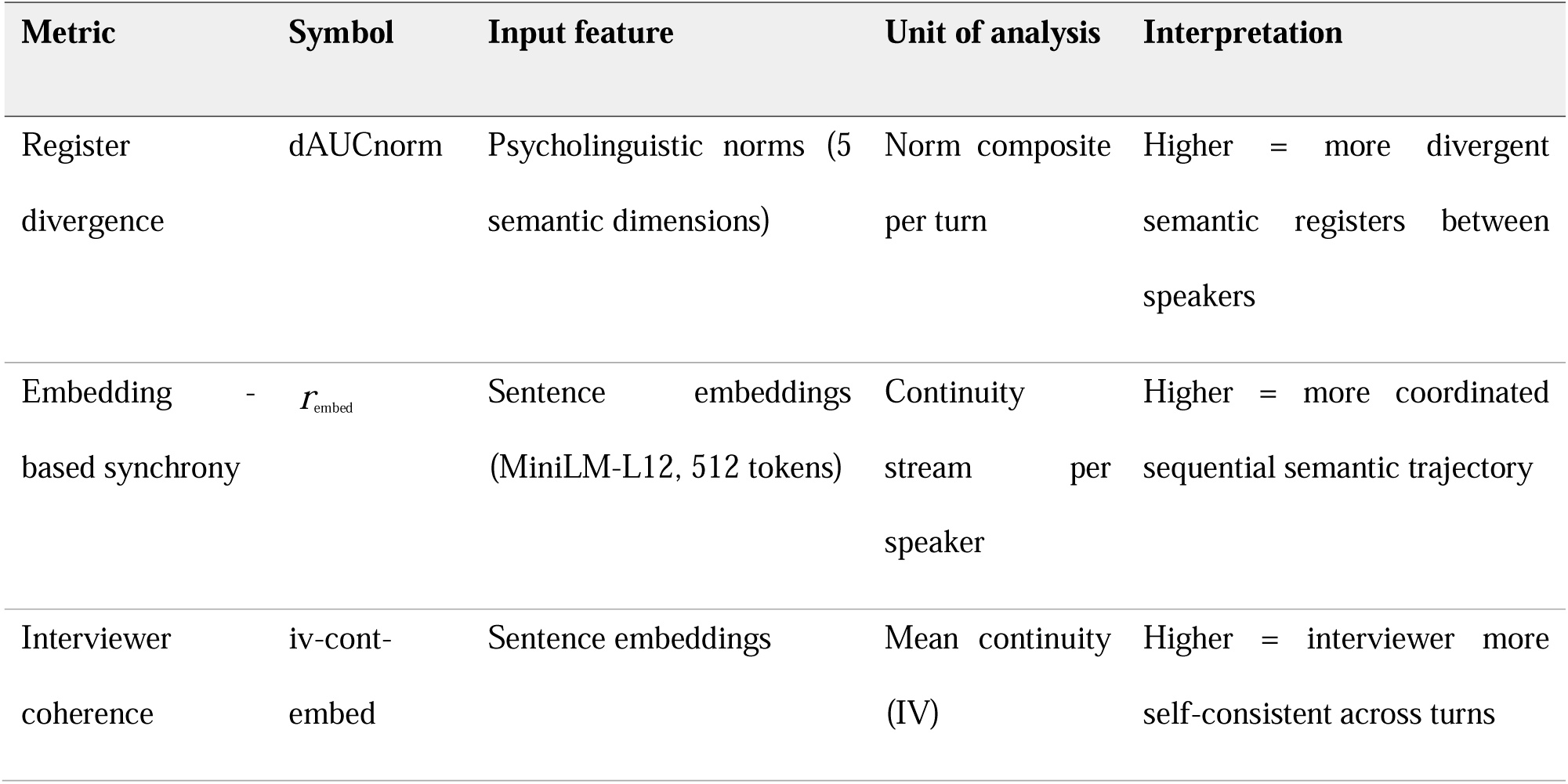
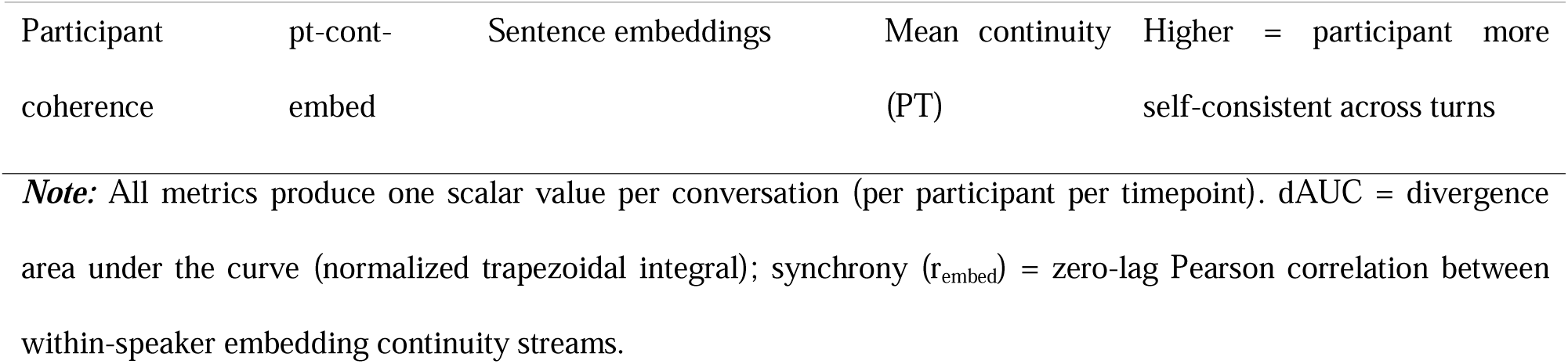
Summary of the primary alignment metrics.

### SM7. Sensitivity analysis for interviewer identity

The first six chronologically conducted DISCOURSE interviews were administered by a different interviewer from the interviewer who conducted the remainder of the interviews. These interviews corresponded to baseline assessments, and all involved healthy-control participants. Interviewer identity was therefore confounded with chronology, group, and timepoint within this initial block. Consequently, an interviewer coefficient could not be interpreted as an independent interviewer effect, and the presence of only two interviewers, one represented by six interviews, did not support estimation of interviewer identity as a random effect.

We conducted a deletion sensitivity analysis to determine whether the principal findings depended materially on these six interviews. The five principal conversation-level outcomes were analysed: register divergence (dAUC_norm_), embedding-based divergence (dAUC_embed_), embedding-based synchrony (r_embed_), interviewer within-speaker coherence, and participant within-speaker coherence. For each outcome, we refitted the pooled patient–control model:

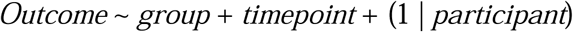

and the group-by-timepoint model:

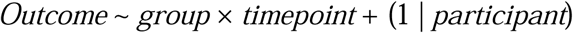

Only the six baseline records were excluded. Available 12-month observations from the same participants were retained because they were conducted by the second interviewer. The full analysis dataset contained 271 conversations from 158 participants. The sensitivity dataset contained 265 conversations from 157 participants: 51 healthy-control and 99 patient interview at baseline, and 45 healthy-control and 70 patient interviews at 12 months.

## Supplementary Results

### SR1: Temporal Trajectory of Register Divergence

**Figure SF1.**
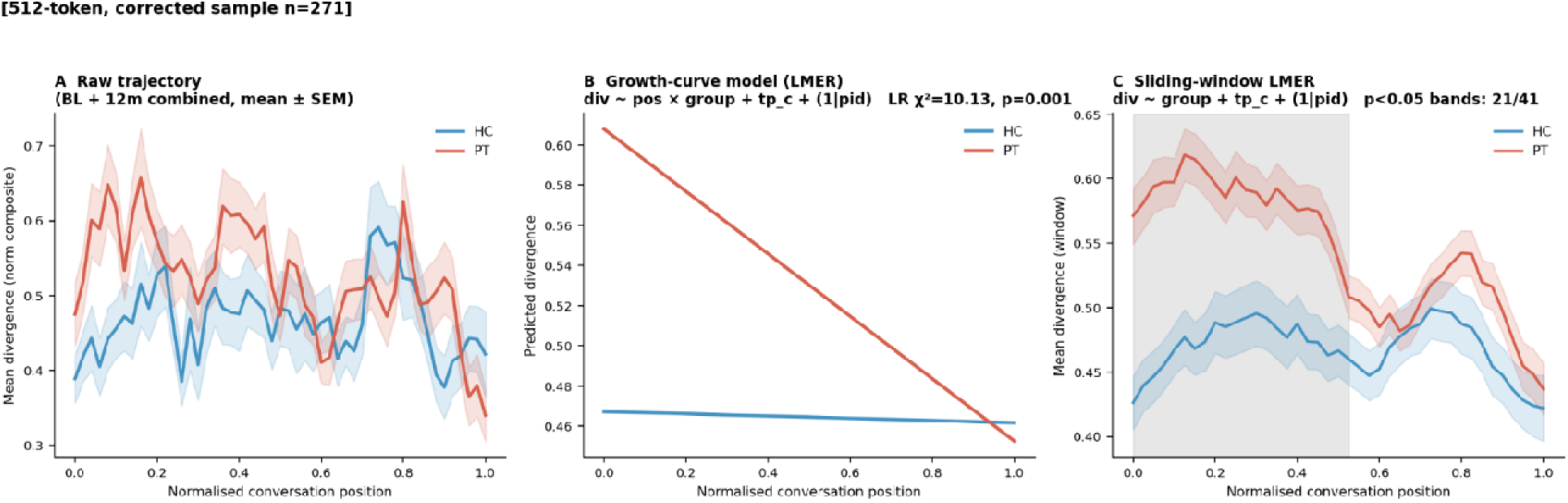
Temporal dynamics of register alignment across the interview (Baseline + 12m combined, n = 271 conversations). Each conversation turn-pair was assigned a normalized position *t*/(n – 1) ∈ [0, 1], where *n* is the total number of paired interviewer–participant exchanges. Divergence at each turn-pair is defined as the absolute difference in per-turn semantic norm composite values (|norm_IV – norm_PT|). Baseline and 12-month conversations pooled; timepoint as a covariate; mixed-model analyses. **Panel A:** Raw mean divergence trajectories (± SEM, interpolated to 51 equally-spaced grid points) separately for healthy controls (HC, blue) and patients (PT, red). **Panel B:** Marginal predictions from a growth-curve mixed-effects model (div ~ pos × group + tp_c + (1|pid)), with a likelihood-ratio test against a model without the position × group interaction (LR χ^2^ = 10.13, df = 1, *p* = .001). **Panel C**: A sliding-window LMER applied at 41 equally-spaced centre points (window half-width = 0.12): at each window, div ~ group_bin + tp_c + (1|pid) was fitted and the group *p*-value extracted; shaded bands mark windows reaching *p* < .05 (21 of 41 windows). HC = healthy controls; PT = patients; tp_c = timepoint contrast-coded (Baseline = –0.5, 12m = +0.5).

Per-turn semantic norm composites were computed for each interviewer and participant turn using the five-dimension psycholinguistic lookup table (ConversationAlign package, Reilly et al.) with each dimension pre-normalized to [0, 1] and the composite for a given turn being the mean of all dimensions with available values across the unique content words in that turn (after lemmatisation and stopword removal). For each conversation, turn-pairs were assigned a normalized position *t*/(n – 1), where *t* is the sequential paired-exchange index (0-based) and *n* the total number of complete paired exchanges (minimum *n* = 5 required for inclusion). Turn-level divergence was |norm_IV(t) – norm_PT(t)|.

Three analyses were conducted on the resulting dataset of 5,486 turn-pair records across 271 conversations. First, raw group trajectories were obtained by interpolating each participant’s divergence values onto a 51-point grid and averaging across participants separately for HC and PT, with bootstrapped SEM. Second, a growth-curve linear mixed-effects model (div ~ pos × group_bin + tp_c + (1|pid); fitted via statsmodels v0.14) tested whether the conversation-position slope differed between groups, using a likelihood-ratio test against a reduced model without the interaction term. Third, a sliding-window LMER (div ~ group_bin + tp_c + (1|pid)) was applied at 41 equally-spaced positions across [0, 1] (window half-width = 0.12), extracting the group β coefficient and *p*-value at each window. No multiple-comparison correction was applied to the window-level *p*-values; windows are reported descriptively to characterize the spatial distribution of the group effect across conversation structure.

Patients showed consistently higher norm divergence from the interviewer than healthy controls across the full length of the interview, indicating that register dissimilarity is a pervasive rather than transient feature of schizophrenia-spectrum speech. The significant position × group interaction in the growth-curve model (LR χ^2^ = 10.13, *p* = .001) indicates that this gap is not static: the PT-HC divergence was greatest during earlier, less scaffolded portions of the protocol and attenuated as later sections introduced stronger visual or textual anchors. Because divergence was defined as an absolute interviewer-participant distance, this analysis does not identify whether attenuation reflected patient convergence, interviewer accommodation, or reciprocal adaptation.

**Figure SF2:**
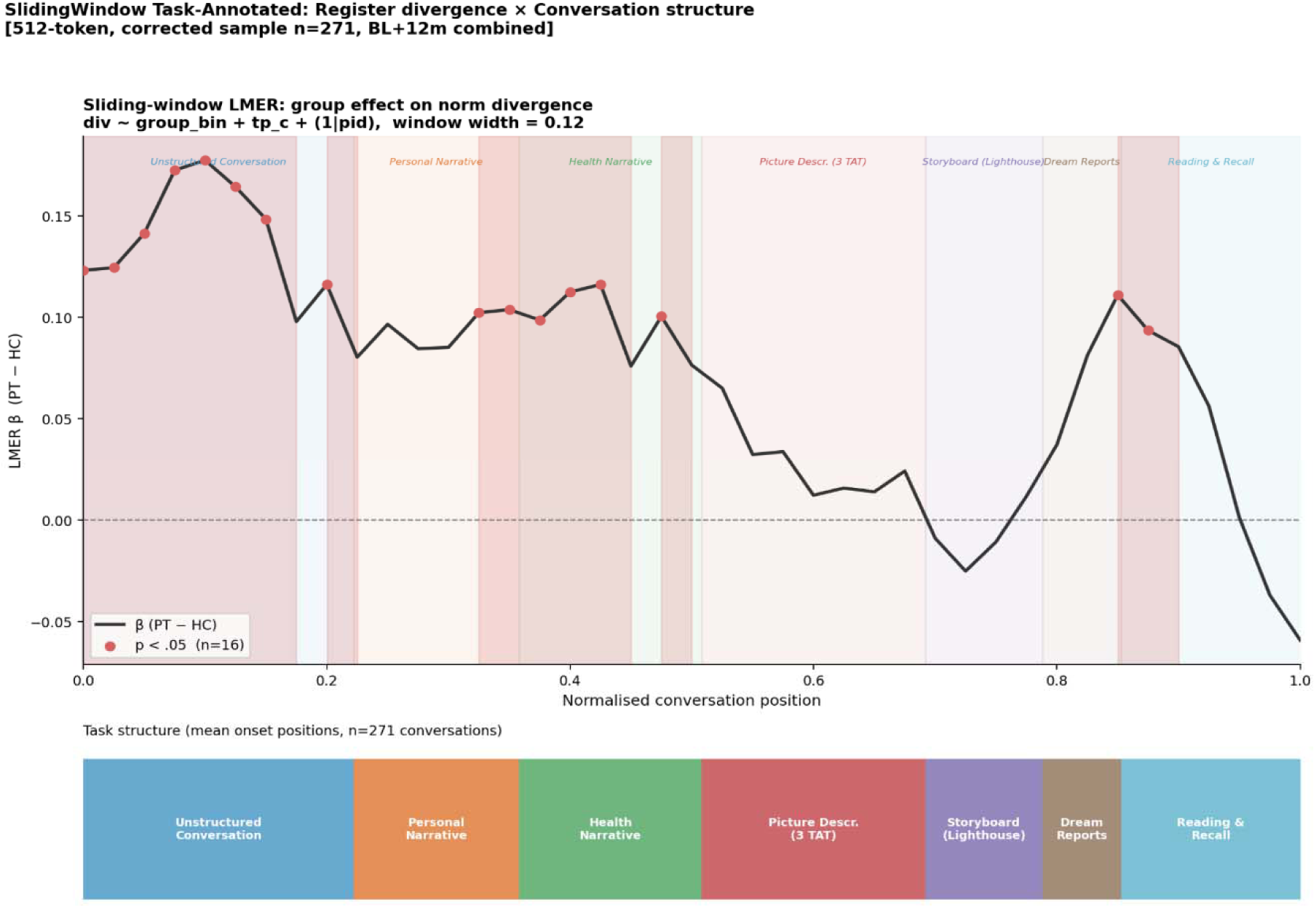
Task-dependent nature of register divergence. The diagnostic group effect for register divergence is the largest and most sustained during the opening Unstructured Conversation segment (normalized position ≈ 0.00–0.20), where expressive language is least constrained by external task demands. A secondary peak appears aroun Dream Reports (around 0.79–0.85), which also places high demands on self-generated narrative. In contrast, the group effect attenuates markedly during the more guided, external referent-based Picture Description (TAT) and Storyboard tasks (≈ 0.50–0.79). These descriptive patterns show that patient–control differences were largest in open-ended, spontaneous speech contexts and smaller during later externally scaffolded sections. Because task order was fixed, the contributions of discourse context, external scaffolding, conversation position, and increasing familiarity cannot be separated.

### SR2: Dimension Decomposition

To identify which lexical norm dimensions drive the HC–PT group difference in dAUC_norm_, per-turn divergences were computed separately for each of the five constituent dimensions: Auditory (sem_auditory_rescale), Concreteness (sem_cnc_b24_rescale), Contextual Diversity (sem_diversity_rescale), Semantic Neighbours (sem_neighbors_rescale), and Visual (sem_visual_rescale). The same sliding-window LMER procedure as the main temporal analysis was applied (41 equally-spaced windows, half-width = 0.12): divergence_dim ~ group_bin + tp_c + (1|participant_ID). The number of windows reaching p < .05 served as an index of the dimension’s discriminative breadth across the conversation.

**Table ST3.**
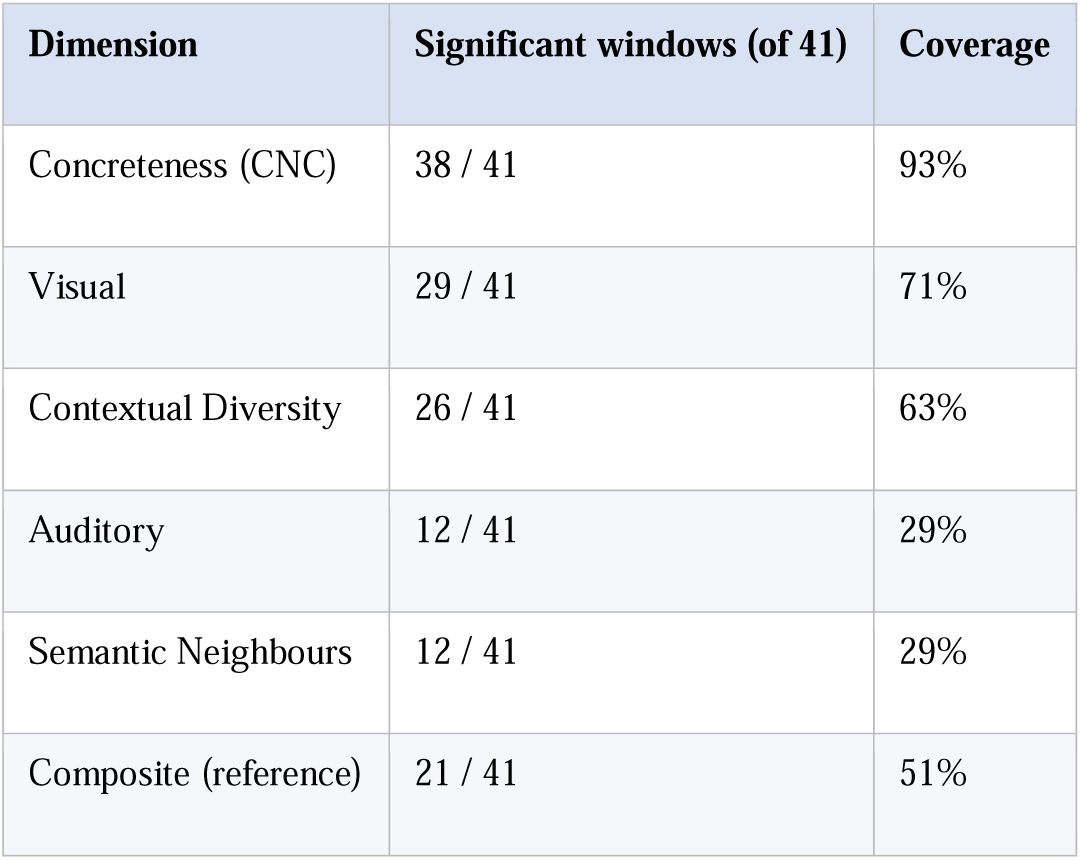
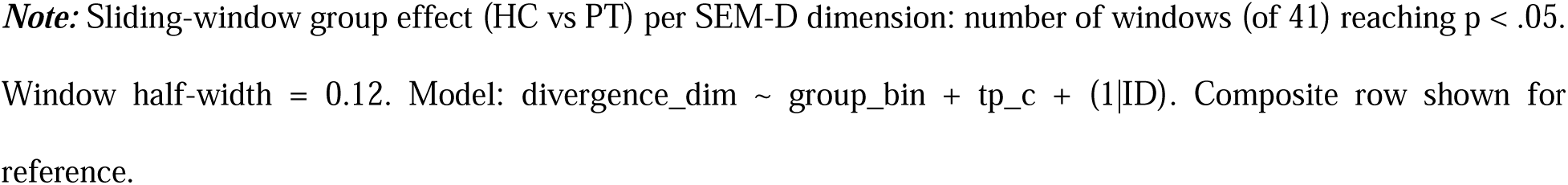
Sliding Window Dimension Decomposition.

| Dimension | Significant windows (of 41) | Coverage |
| --- | --- | --- |
| Concreteness (CNC) | 38 / 41 | 93% |
| Visual | 29 / 41 | 71% |
| Contextual Diversity | 26 / 41 | 63% |
| Auditory | 12 / 41 | 29% |
| Semantic Neighbours | 12 / 41 | 29% |
| Composite (reference) | 21 / 41 | 51% |

**Note:** Sliding-window group effect (HC vs PT) per SEM-D dimension: number of windows (of 41) reaching $p < .05$ . Window half-width = 0.12. Model: $\text{divergence\_dim} \sim \text{group\_bin} + \text{tp\_c} + (1|\text{ID})$ . Composite row shown for reference.

Concreteness was by far the most discriminative dimension: 38 of 41 windows reached significance, compared with 21 for the composite. Visual and Contextual Diversity also showed broad group separation (71% and 63% of windows, respectively). Auditory and Semantic Neighbours were less discriminative (29% each). This decomposition is consistent with reduced use of embodied, imageable vocabulary in patient speech. The composite signal thus reflects primarily the concreteness and visual contributions.

**Figure SF3:**
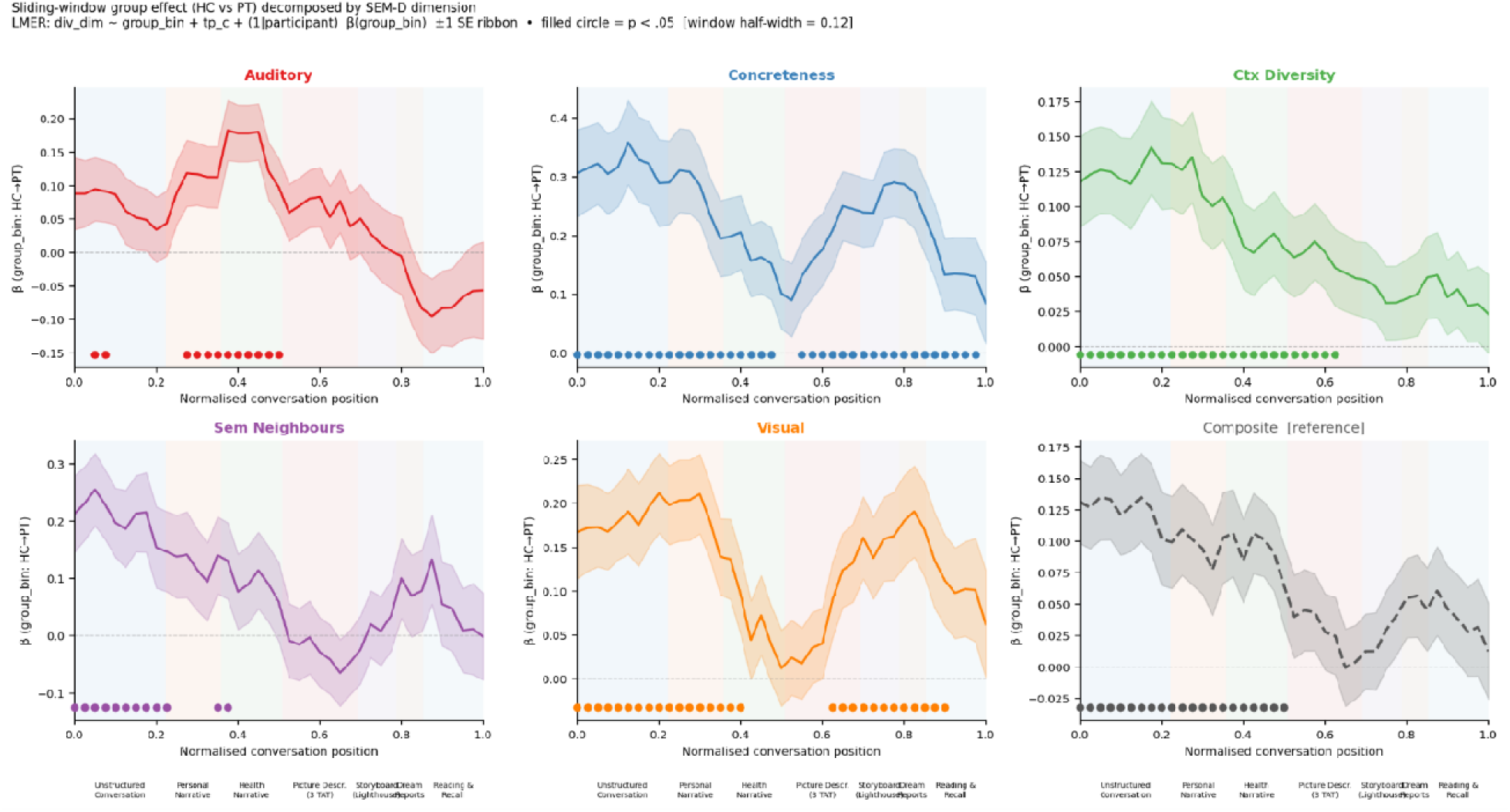
Sliding windows for individual psycho-linguistic dimensions. ***Note:*** Sliding-window group effect (HC vs PT) decomposed by lexical norm dimension. Each panel shows beta(group_bin) ± 1 SE across normalized conversation position (0 = start, 1 = end). Filled circles indicate windows with p < .05. Task bands reflect mean onset positions of seven structured conversation segments across all 271 transcripts.

Plots show individual trajectories across normalized conversation positions for each dimension, annotated by task segment. Similar to the composite signal, where register divergence was seen mostly in the first half, individual dimensions also showed maximal HC/PT differences in the early part of the interview.

### SR3: Embedding register divergence (dAUC_embed_)

**Fig SF4:**
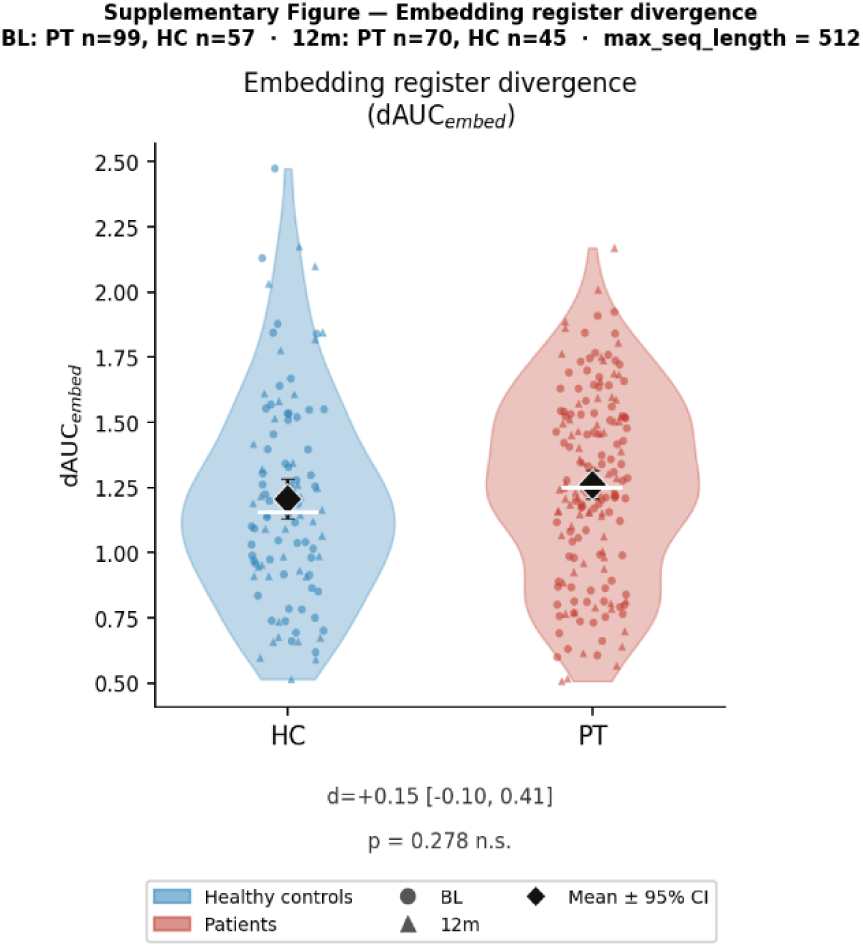
Turn-by-turn divergence of embedding-based register (dAUC_embed_): Unlike the norm-based dAUC, turn-by-turn divergence of embedding-based register (dAUC_embed_) did not differ significantly between patients and healthy controls (Cohen’s d = +0.15 [−0.10, 0.41], p = .278). dAUC_embed_ is derived from PCA on joint IV–PT sentence embeddings per conversation, and the first principal component captures the dominant direction of within-dyad semantic variation (i.e., the single axis that best summarises how meaning shifts across all turns in *that specific conversation)*. Because the embedding space integrates syntactic and contextual information alongside semantic content, it is less sensitive than the norm composite to the specific vocabulary-level register differences that characterize impoverished speech. The contrast between a significant dAUC_norm_ and a null dAUC_embed_ effect supports the view that the norm pipeline selectively captures register differences at the level of semantic content richness, while the embedding pipeline is relatively insensitive to this dimension, consistent with the two metrics serving complementary rather than redundant roles.

### SR4: Inter-correlation among alignment metrics

**Figure SF5:**
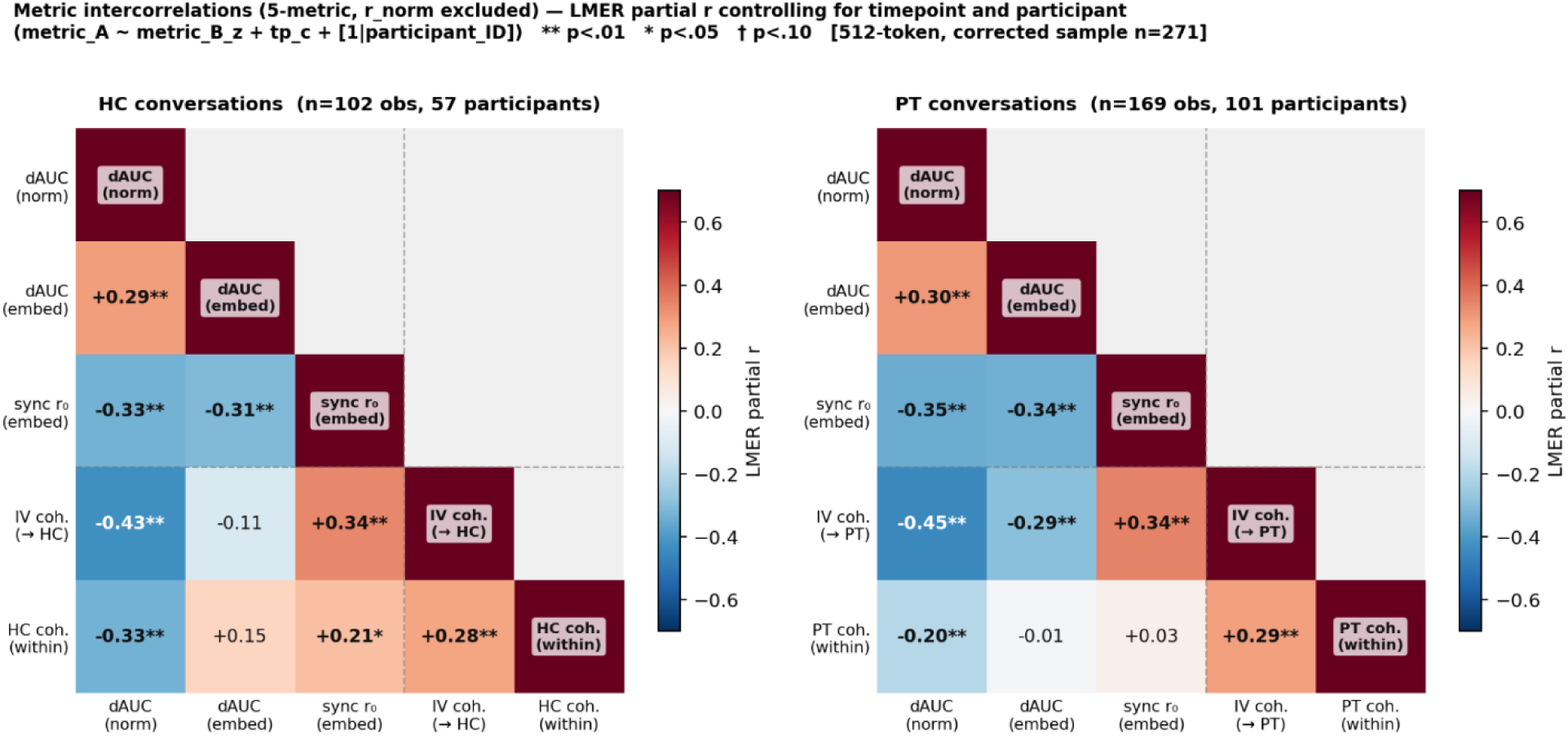
Alignment metric intercorrelations: Heatmaps from conversations involving healthy controls (left) are shown separately from patients (right). Examining the inter-correlations among the alignment matrix reveals that higher divergence in psycholinguistic register from the participant is associated with lower embedding-based synchrony and reduced within-person coherence in both parties (interviewers and participants). On the other hand, higher embedding-based conversational synchrony tracks higher within-person coherence in interviewers and healthy controls (but not patients). *dAUC* = divergence Area Under the Curve, computed as the trapezoidal integral of |IV_score(t) − PT_score(t)| across paired turns, normalized by n − 1 turn-pairs; *norm* subscript = metric derived from the 5-dimension psycholinguistic semantic composite. *embed* subscript = metric derived from turn-level sentence embeddings (paraphrase-multilingual-MiniLM-L12-v2; max_seq_length = 512). r_embed_ zero-lag Pearson r between the within-speaker embedding continuity streams of interviewer and participant (continuity at turn t = cosine similarity between consecutive turn embeddings). *IV coh.* = mean within-speaker turn-to-turn cosine similarity of the interviewer, stratified by the conversation partner’s diagnostic group (→ HC: interviews with healthy controls; → PT: interviews with patients). *HC/PT coh. (within)* = mean within-speaker turn-to-turn cosine similarity of the participant (healthy control or patient, respectively). Cells show LMER partial r from the model: metric_A ~ metric_B_z + tp_c + (1 | participant_ID), where metric_B_z is the z-scored predictor, tp_c is timepoint contrast-coded (baseline = −0.5, 12-month = +0.5), and (1 | participant_ID) is a random intercept for participants. ** p < .01; * p < .05; † p < .10. Sample: n = 271 conversations (BL = 156, 12m = 115); HC panel n = 102 observations, 57 participants; PT panel n = 169 observations, 101 participants.

The dissociation between the norm-based and embedding-based metrics has methodological and theoretical implications. The null finding for embedding-based register divergence contrasts sharply with the significant norm-based effect, confirming that sentence-embedding space integrates syntactic and contextual information in a way that dilutes the lexical-level register differences that characterize impoverished speech. The two metric families are therefore complementary: norms capturing the register anchored in habitual vocabulary and embeddings capturing the sequential dynamics of meaning-making. The inter-metric correlation analysis further confirms this complementarity: within patients, register divergence and synchrony are negatively correlated, but neither maps straightforwardly onto within-speaker coherence, suggesting three partially independent loci of communicative breakdown.

### SR5: Severity of thought disorder and conversational alignment

**Figure SF6:**
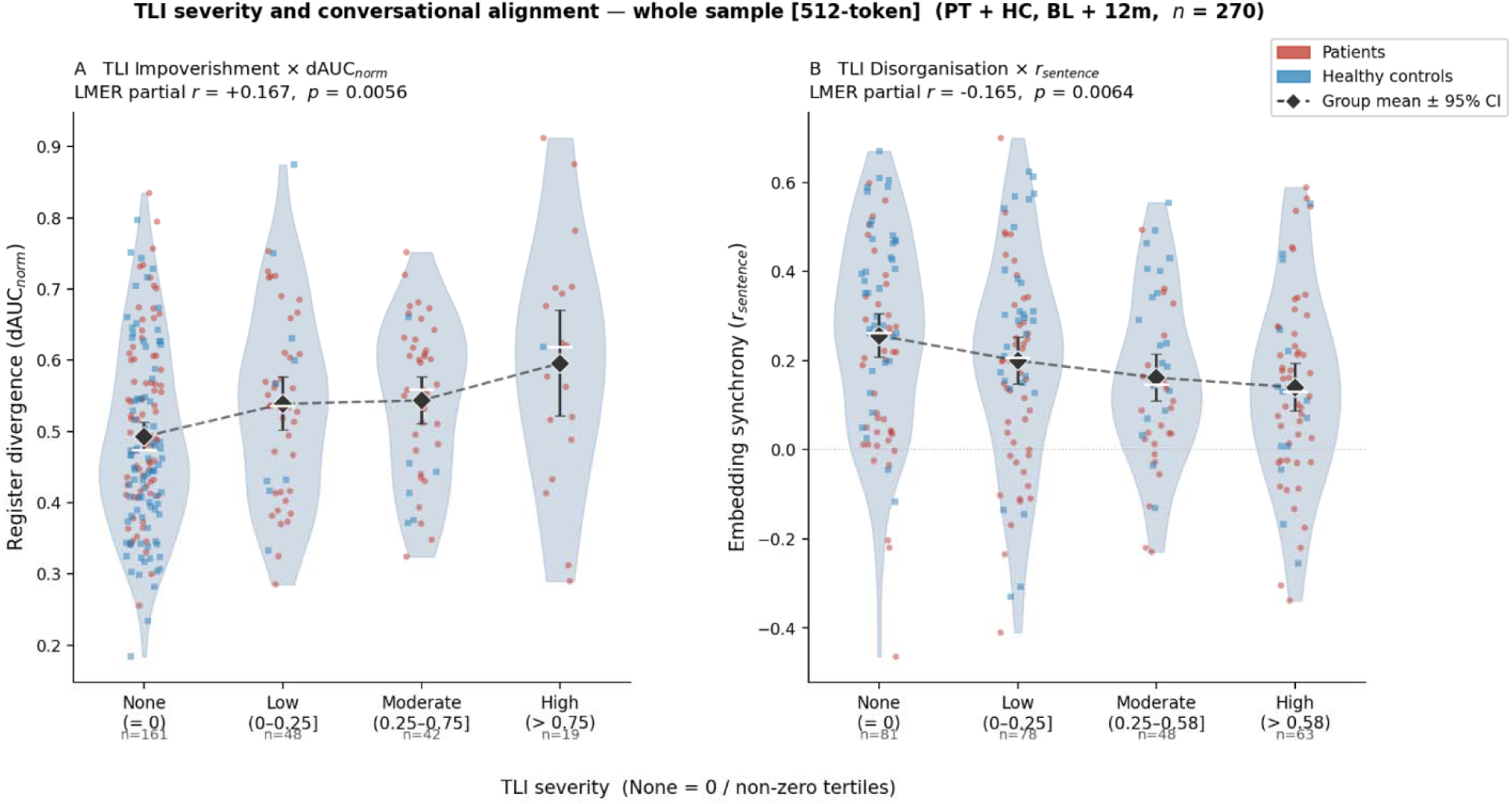
Register divergence and embedding synchrony by TLI severity bin. TLI severity was divided into four bins: None (score = 0), Low (0–0.25), Moderate (0.25–0.75 / 0.58 for Disorganization), and High (> 0.75 / > 0.58), derived from non-zero tertiles within the patient group. Diamonds and error bars show the group means ± 95% CI. Dashed lines show the LMER partial r trend. Patients are shown in red, healthy controls in blue. PT + HC, BL + 12m combined; n = 270; dAUC_norm_; Panel A; r_embed_; Panel B. LMER partial r values control for timepoint and participant (metric ~ TLI_z + tp_c + [1|participant_ID]). TLI Impoverishment: partial r = +0.167, p = .0056; TLI Disorganization: partial r = −0.165, p = .0064. Severity bins are shown for visualization only; inferential tests used continuous TLI scores. An additional group- and timepoint-adjusted sensitivity analysis of register divergence an TLI Impoverishment is reported in Supplementary Results SR5.

The reported primary continuous-score analyses showed associations of TLI Impoverishment with greater register divergence and TLI Disorganization with reduced embedding-based synchrony (both BH-adjusted q = .038; Figure 3). Mean register divergence also increased across the visualised TLI Impoverishment severity bins, although inference was based on continuous scores rather than the binned categories.

To test whether the impoverishment association was driven by the small High-severity bin, we conducted additional robustness analyses on the archived per-conversation metric values. Leave-one-participant-out refits (157 per model) produced no sign reversal, and the association remained significant after every deletion (primary model β range +0.028 to +0.038; group-adjusted model +0.021 to +0.031). Excluding all patient observations in the High bin (TLI Impoverishment > 0.75) left the association intact under the primary model (β = +0.040, 95% CI [+0.005, +0.076], p = .026) and attenuated it under the group-adjusted model (p = .162); this exclusion removes 74% of the predictor variance among patients (SD 0.51 to 0.26), so attenuation under the group-adjusted specification is the expected consequence of range restriction. Two analyses that retain all observations while capping the influence of extreme scores remained significant under both specifications: rank transformation of the predictor (primary p = .0009; group-adjusted p = .026) and winsorization at the High-bin threshold (primary p = .002; group-adjusted p = .035). The association is therefore graded across the severity range rather than carried by its upper end.

### SR6: Conversational floor structure in patients and controls

Floor occupation was operationalized as each speaker’s proportional share of total conversation. Per-conversation metrics were: participant floor in word count (%), interviewer floor (%), total participant words, total turns (both speakers combined), and mean participant turn length (words). Each metric was modelled with LMER including Group (PT vs HC), Timepoint (BL vs 12m), and their interaction as fixed effects and random intercepts for participant ID.

**Table ST4:** Group effects for conversational floor metrics.

| Metric | HC Mean (SD) | PT Mean (SD) | Group *r* | *p* | G×T *r* | *p* |
| --- | --- | --- | --- | --- | --- | --- |
| Participant floor (%) | 74.9 (5.8) | 67.0 (11.0) | -0.310 | < .001*** | +0.032 | .605 |
| Interviewer floor (%) | 25.1 (5.8) | 33.0 (11.0) | +0.310 | < .001*** | -0.032 | .605 |
| Participant total words | 2126 (627) | 1831 (685) | -0.183 | .002** | -0.123 | .042* |
| Total turns | 52.4 (22.9) | 75.9 (35.1) | +0.319 | < .001*** | -0.091 | .135 |
| Mean turn length (words) | 92.6 (36.3) | 61.1 (37.5) | -0.324 | < .001*** | +0.036 | .550 |

**Notes:** Floor metrics: mean (SD) collapsed across timepoints and LMER group effects. Model: metric ~ Group + Timepoint + Group×Timepoint + (1|ID). HC n = 57 participants (102 obs); PT n = 99–101 participants (169 obs). G×T = Group×Timepoint interaction. \*\*\* $p < .001$ , \*\* $p < .01$ , \* $p < .05$ .

Patients occupied a significantly smaller share of the conversation with fewer total words and a fragmented conversational structure characterised by more numerous but briefer exchanges. These structural differences were stable across BL and 12m (all G×T interactions p > .13; see Table ST4), except for participant total words where a diverging trajectory emerged: HC increased word output from BL to 12m while PT remained stable (interaction r = −0.12, p = .042).

**Figure SF7:**
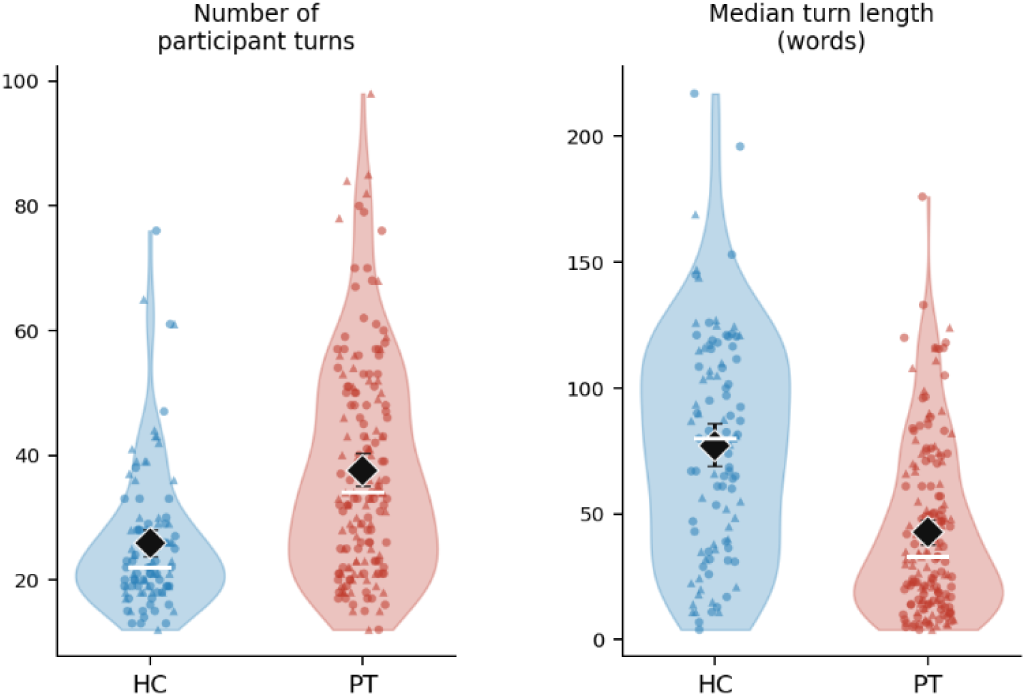
Number and length of conversational turns in patients and controls. *Notes:* HC: Healthy controls PT: Patient. 12m data in triangles, baseline in dots.

### SR7: Reliability-Weighted dAUC and r_embed_

The standard dAUC_norm_ is a normalized trapezoidal integral of |IV_nc(t) − PT_nc(t)| across sequential turn-pairs, where _nc denotes the mean semantic composite (across five dimensions) over matched content words, and given this averaging, it is implicitly length-normalized at the word level. However, a mean over shorter turns (e.g., two matched words) carries higher sampling variance than longer turns (e.g., twenty words), leading to noisier turn-pair estimates that may inflate |IV_nc − PT_nc| register divergence.

To test this, each turn-pair t was assigned a reliability weight equal to the harmonic mean of matched-word counts for interviewer (n_iv) and participant (n_pt) turns:

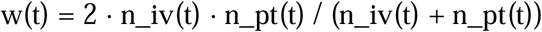

The weighted metric was then computed as dAUC_norm__w = Σ(w_t · |IV_nc_t − PT_nc_t|) / Σ(w_t). Three evaluations were conducted: (1) HC–PT group separation, (2) within-group floor sensitivity; i.e., whether the correlation between dAUC and median participant turn length attenuates under weighting and (3) TLI clinical associations in the patient group.

An analogous reliability-weighting was applied to *r*_embed_(zero-lag Pearson correlation between the interviewer and participant continuity streams). Each continuity step *t* is a cosine similarity between consecutive same-speaker embeddings i.e., cosine(embed, embed). Its sampling variance scales with the quality of both embeddings, which degrades for shorter turns. Reliability weights for each step were therefore defined as the harmonic mean of adjacent-turn word counts within each speaker, and the joint weight for position *t* as the geometric mean of the two speaker weights:

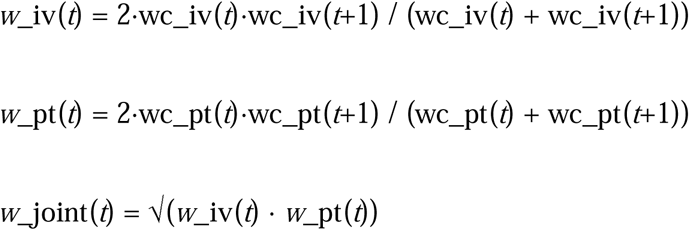

The weighted metric *r*_embed__w is the weighted Pearson correlation of the two continuity streams with weights *w*_joint. Mean harmonic weights per conversation were HC: 28.3 (SD 12.4) vs PT: 20.9 (SD 12.0), consistent with the shorter participant turns established in Section 1.

**Table ST5:**
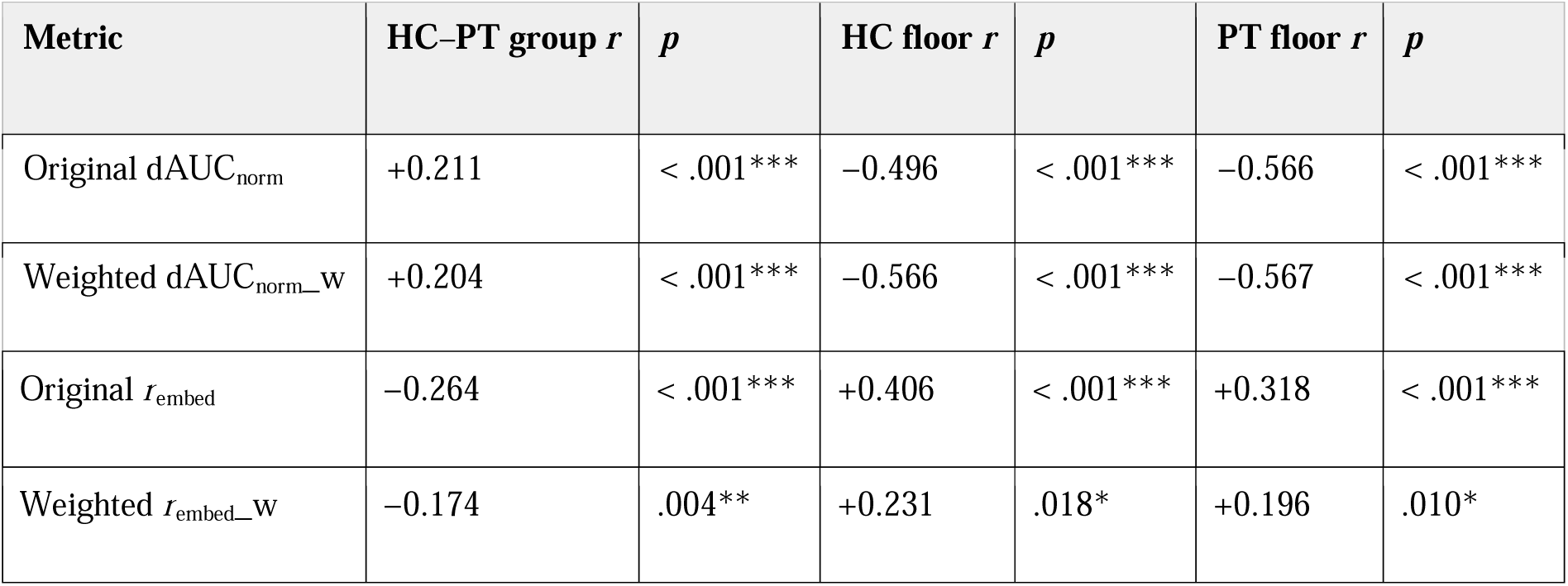
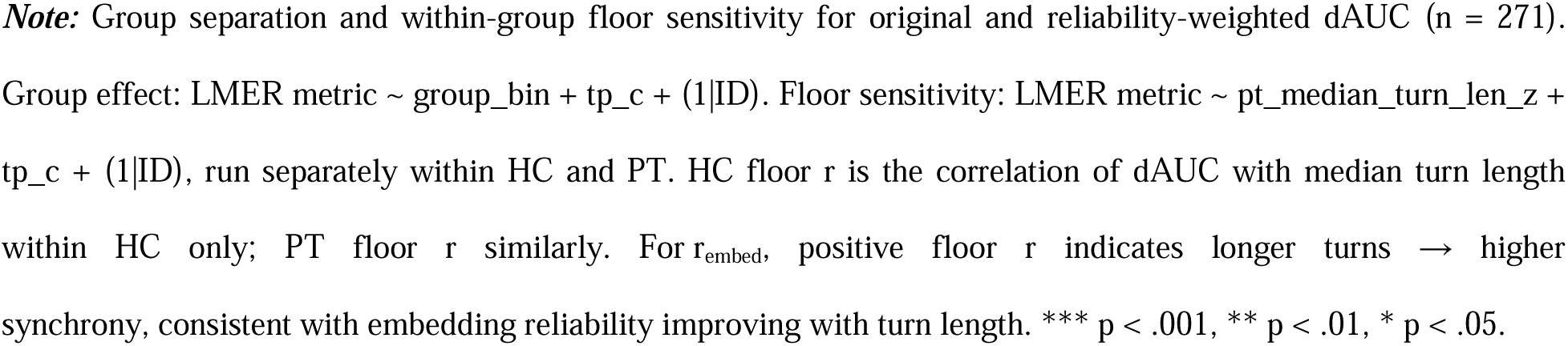
Group separation and floor sensitivity for the original and weighted metrics.

| Metric | HC-PT group $r$ | $p$ | HC floor $r$ | $p$ | PT floor $r$ | $p$ |
| --- | --- | --- | --- | --- | --- | --- |
| Original $\text{dAUC}_{\text{norm}}$ | +0.211 | < .001*** | -0.496 | < .001*** | -0.566 | < .001*** |
| Weighted $\text{dAUC}_{\text{norm\_w}}$ | +0.204 | < .001*** | -0.566 | < .001*** | -0.567 | < .001*** |
| Original $r_{\text{embed}}$ | -0.264 | < .001*** | +0.406 | < .001*** | +0.318 | < .001*** |
| Weighted $r_{\text{embed\_w}}$ | -0.174 | .004** | +0.231 | .018* | +0.196 | .010* |

**Note:** Group separation and within-group floor sensitivity for original and reliability-weighted dAUC ( $n = 271$ ). Group effect: LMER metric ∼ group\_bin + tp\_c + (1|ID). Floor sensitivity: LMER metric ∼ pt\_median\_turn\_len\_z + tp\_c + (1|ID), run separately within HC and PT. HC floor $r$ is the correlation of dAUC with median turn length within HC only; PT floor $r$ similarly. For $r_{\text{embed}}$ , positive floor $r$ indicates longer turns → higher synchrony, consistent with embedding reliability improving with turn length. \*\*\* $p < .001$ , \*\* $p < .01$ , \* $p < .05$ .

Reliability weighting left the dAUC_norm_ group effect essentially unchanged (+0.211 vs +0.204), and floor sensitivity was not attenuated; within HC it became marginally stronger (−0.496 vs −0.566). Near-identical floor correlations across groups (both *r* ≈ −0.57 after weighting) confirm that this relationship is a general compositional property of the metric: longer turns yield more matched content words and systematically higher composite values, reducing the norm divergence regardless of group. Reliability weighting is therefore not corrective for this floor sensitivity but sharpens sensitivity to genuine clinical signal by suppressing high-variance turn-pairs.

The pattern for *r*_embed_ was qualitatively different. The HC–PT group effect attenuated under weighting (−0.264 → −0.174), reflecting the fact that HC conversations carry substantially higher mean harmonic weights than PT conversations (28.3 vs 20.9 words per step): down-weighting short-turn steps reduces the HC advantage in a way that is not purely compositional. Floor sensitivity was partially but not fully eliminated: the correlation of *r*_embed_ with median participant turn length was reduced by approximately 40–45% in both groups (HC: +0.406 → +0.231; PT: +0.318 → +0.196) but remained significant. This indicates that the turn-length dependence of *r*_embed_ has two components: one attributable to embedding noise in short turns (removed by weighting) and one structural, reflecting that more numerous, shorter turns create more within-speaker discontinuities in the continuity stream regardless of embedding quality.

**Table ST6:** TLI Clinical Associations (PT only, n = 167)

| Predictor → Outcome | partial $r$ | $p$ |
| --- | --- | --- |
| TLI Impoverishment → original $\text{dAUC}_{\text{norm}}$ | +0.201 | .009** |
| TLI Impoverishment → weighted $\text{dAUC}_{\text{norm\_w}}$ | +0.327 | < .001*** |
| TLI Disorganization → original $\text{dAUC}_{\text{norm}}$ | -0.115 | .138 |
| TLI Disorganization → weighted $\text{dAUC}_{\text{norm\_w}}$ | -0.120 | .123 |
| TLI Impoverishment → original $r_{\text{embed}}$ | -0.111 | .151 |
| TLI Impoverishment → weighted $r_{\text{embed\_w}}$ | -0.102 | .189 |
| TLI Disorganization → original $r_{\text{embed}}$ | -0.156 | .043* |
| TLI Disorganization → weighted $r_{\text{embed\_w}}$ | -0.179 | .020* |

**Note:** LMER: metric ~ predictor\_z + tp\_c + (1|ID). Adjusted models add n\_turns\_z as covariate. \*\*\* $p < .001$ , \*\* $p < .01$ , \* $p < .05$ , † $p < .10$ .

Reliability weighting substantially sharpened the TLI Impoverishment association for dAUC_norm_ (*r* = +0.20 vs +0.33), consistent with impoverished speakers contributing more low-weight, high-noise turn-pairs that dilute clinical signal in the unweighted metric. After controlling for total number of turns, dAUC_norm__w retained a significant independent association with TLI Impoverishment (*r* = +0.178, *p* = .021), while turn count was not independently significant (*r* = +0.141, *p* = .069), confirming that the weighted metric captures genuine semantic divergence beyond the contribution of conversation structure. For *r*_embed_, the pattern was complementary. Neither the original nor the weighted metric was associated with TLI Impoverishment (both *r* ~ −0.10, *p* > .15), and both collapsed entirely when number of turns was added as a covariate, indicating that any apparent synchrony deficit in more impoverished patients is fully mediated by conversational fragmentation. TLI Disorganization, by contrast, showed a significant association with *r*_embed__w (*r* = −0.179, *p* = .020) that was absent for the unweighted version in the adjusted model: after controlling for turn count, *r*_embed__w remained significant (*r* = −0.173, *p* = .025) while the unweighted *r*_embed_ dropped to a trend (*r* = −0.149, *p* = .054). This indicates that reliability weighting recovers a specific association between reduced dyadic continuity synchrony and positive formal thought disorder that is masked in the unweighted metric by noise from short-turn steps.

The two weighted metrics thus capture dissociable dimensions of thought disorder: dAUC_norm__w indexes poverty of speech through increased dyadic semantic *divergence*, while *r*_embed_-w indexes disorganization through reduced *synchrony* of the speakers’ semantic trajectories. Both associations are independent of turn count structure once the appropriate weighting is applied.

### SR8: 128- vs. 512-token embeddings

As healthy controls produced substantially longer turns than patients (Supplementary Table ST4), embeddings generated using longer inputs may bias the HC vs patient group difference in *r*_embed_ and within-speaker coherence metrics. To address this, we truncated input tokens per turn to 128-token limit and recomputed embeddings. Table ST7 presents the 128-token results. All primary inferential conclusions were preserved. Participant coherence remained non-significant, although its point estimate changed sign. The principal quantitative difference was a somewhat smaller group effect for rembed with 128-token encoding (d = −0.59 at 512 tokens vs −0.50 at 128 tokens).

**Table ST7:** LMER group comparisons for 128-token versus 512-token embeddings.

| Metric | HC mean | PT mean | p (128) | d (128-token) | d (512-token) |
| --- | --- | --- | --- | --- | --- |
| $\text{dAUC}_{\text{norm}}$ | 0.478 | 0.541 | < .001 | +0.48 | +0.48 |
| $r_{\text{embed}}$ | 0.273 | 0.148 | < .001 | -0.50 | -0.59 |
| iv-cont-embed | 0.389 | 0.373 | .017 | -0.33 | -0.33 |
| pt-cont-embed | 0.293 | 0.287 | .374 | +0.12 | -0.10 |

**Note:** LMER group comparison results for 128-token versus 512-token embeddings (Baseline + 12m pooled, $n = 271$ ). $dAUC_{\text{norm}}$ is derived from psycholinguistic norms (not embeddings) and is therefore unaffected by sequence length (replicated identically). For all embedding-based metrics, the inferential conclusion concerning statistical significance was preserved under both truncation limits. Participant coherence remained non-significant, although its point estimate changed sign.

**Figure SF8:**
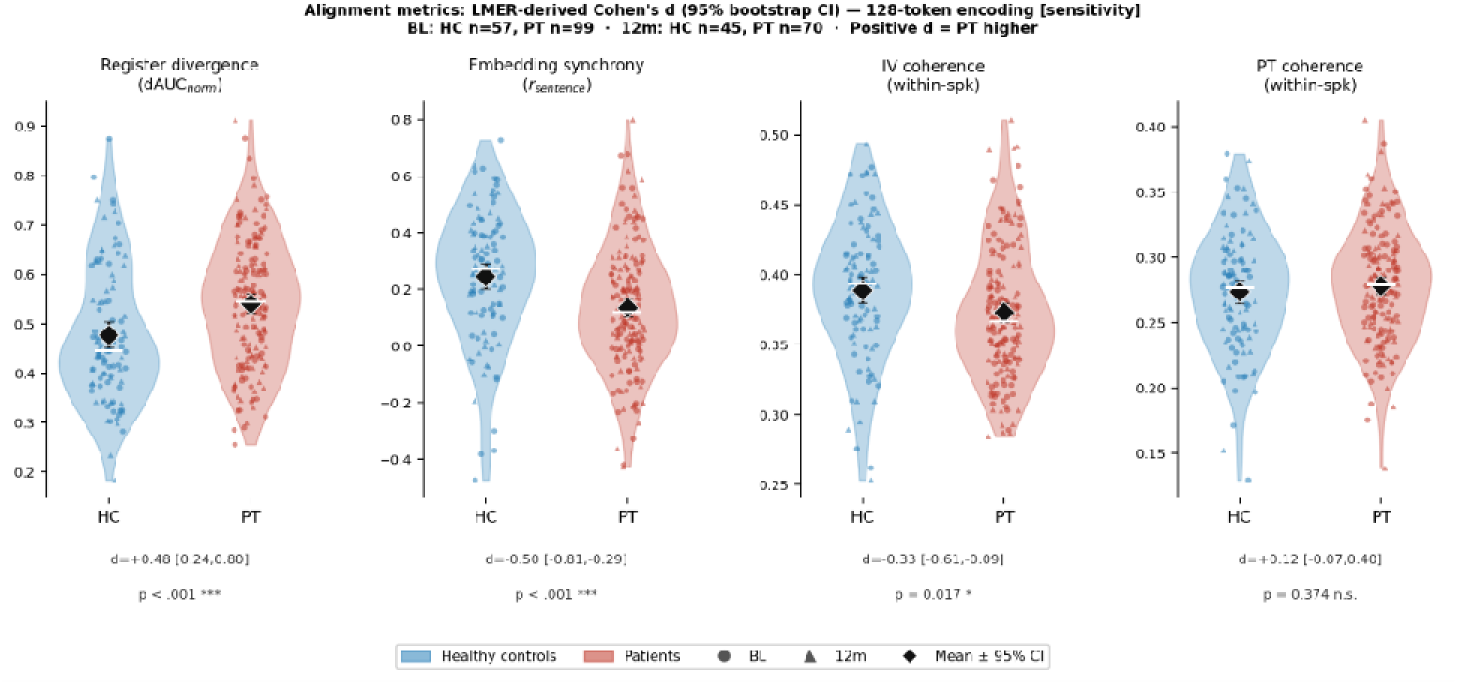
Group differences in conversational alignment metrics with 128 token embeddings. ***Notes***: Violin plots with individual observations and mean markers with 95% confidence intervals showing distributions of each alignment metric separately for healthy controls (HC, blue) and patients (PT, red), at baseline (BL) and 12-month follow-up (12m). Panel A: Register divergence (dAUC_norm_); higher values indicate greater divergence in psycholinguistic register between interviewer and participant. Panel B: Embedding-based rhythmic synchrony (r_embed_); higher values indicate greater coupling of the two speakers’ sequential semantic trajectories. Panel C: Interviewer within-speaker coherence; higher values indicate greater semantic self-consistency of the interviewer across turns. Panel D: Participant within-speaker coherence. LMER group main effect (* p < .05; ** p < .01; *** p < .001); see Table ST7 for full model parameters. HC n = 57 participants (102 observations); PT n = 99 participants (169 observations). Effect sizes are LMER-derived Cohen’s d with 95% bootstrap CI.

**Figure SF9:**
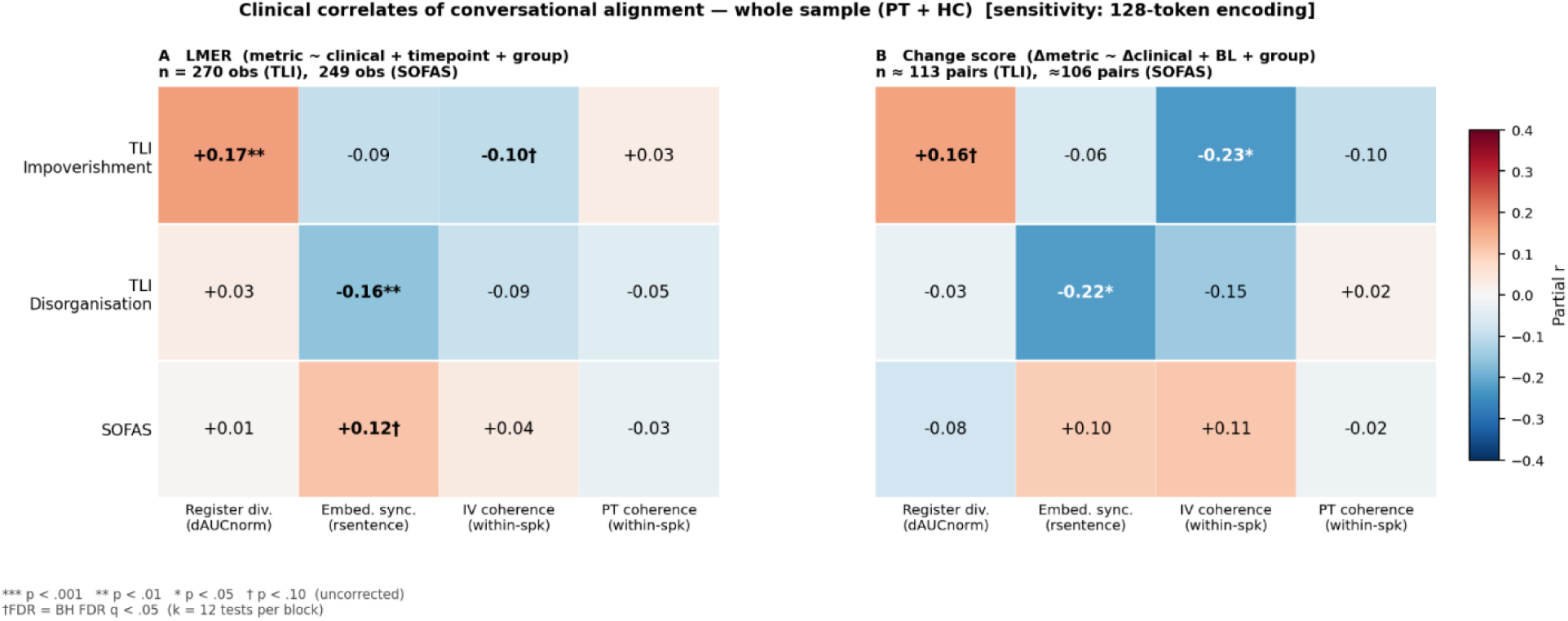
Clinical correlates of conversational alignment *Notes*: Heatmaps show partial r values for four alignment metrics (columns) against three clinical variables (rows). Colour indicates the direction and magnitude of the association (red, positive; blue, negative). Asterisks indicate uncorrected significance: *** p < .001, ** p < .01, and * p < .05; † indicates p < .10. SOFAS = Social and Occupational Functioning Assessment Scale; TLI = Thought and Language Index. Two associations survived FDR correction: dAUC_norm_ and TLI Impoverishment (q = .038), and synchrony (r_embed_) and TLI Disorganization (q = .038).

### SR9: Diagnostic Subgroup Analysis

We examined if alignment metrics differentiated schizophrenia from other early-psychosis diagnoses within the patient group. Within the patient group, schizophrenia (n = 58 participants, 89 observations) was compared with other psychosis diagnoses (Schizoaffective n = 24, Psychosis NOS n = 22, Bipolar with psychotic features n = 5; total 51 participants, 80 observations). Main model: metric ~ SZ + Timepoint + SZ×Timepoint + (1|ID), SZ coded 1 (schizophrenia) vs 0 (other psychosis), with effect sizes reported as partial r = t / √(t^2^ + n − 3). Timepoint was coded BL = −0.5, 12m = +0.5. Embedding-based alignment metrics (r_embed_, dAUC_embed_, within-person interviewer and participant coherence) were computed from 512-token context window (MiniLM-L12-v2). Norm-based analyses (e.g., for the dimension decomposition analysis) operate on lemmatised content-word lookups.

**Table ST8:** Group means and LMER effects across five alignment metrics.

| Metric | SZ Mean (SD) | Other PT Mean (SD) | SZ partial $r$ | $p$ |
| --- | --- | --- | --- | --- |
| dAUC (norm) | 0.570 (0.136) | 0.508 (0.113) | +0.245 | .001** |
| $r_{\text{embed}}$ (embed) | 0.124 (0.187) | 0.174 (0.234) | -0.112 | .146 |
| IV coherence (embed) | 0.365 (0.045) | 0.382 (0.049) | -0.173 | .024* |
| PT coherence (embed) | 0.288 (0.048) | 0.287 (0.051) | +0.011 | .888 |
| $\text{dAUC}_{\text{embed}}$ | 1.312 (0.344) | 1.194 (0.371) | +0.152 | .048* |

**Note:** Alignment metrics by diagnosis (PT only). Means collapsed across timepoints. Main model: $\text{metric} \sim \text{SZ} + \text{Timepoint} + \text{SZ} \times \text{Timepoint} + (1|\text{ID})$ . SZ $n = 89$ obs (58 participants); Other PT $n = 80$ obs (51 participants). All $\text{SZ} \times \text{Timepoint}$ interactions were non-significant (all $p > .80$ ) and are not shown. \*\* $p < .01$ , \* $p < .05$ , † $p < .10$ .

Exploratory diagnosis-subgroup analyses indicated that conversations involving patients with schizophrenia showed greater interviewer–participant divergence than conversations involving other psychotic disorder diagnoses on both dAUC metrics (dAUC_norm_: r = +0.245, p = .001; dAUC_embed_: r = +0.152, p = .048). Interviewer within-speaker coherence was also lower in conversations involving patients with schizophrenia (r = −0.173, p = .024). Interviewers were aware of patient-control status but were not informed of participants’ specific diagnostic subgroup at the time of interview.

**Table ST9:** Subgroup Means by Diagnosis.

| Diagnosis | $n$ obs | dAUC (norm) | $r_{\text{embed}}$ | IV coherence | PT coherence | dAUC (embed) |
| --- | --- | --- | --- | --- | --- | --- |
| Schizophrenia | 89 | 0.571 | 0.124 | 0.365 | 0.288 | 1.312 |
| Schizoaffective Disorder | 39 | 0.487 | 0.207 | 0.381 | 0.280 | 1.162 |
| Psychosis NOS | 32 | 0.532 | 0.142 | 0.386 | 0.291 | 1.180 |
| Bipolar Disorder | 9 | 0.505 | 0.143 | 0.369 | 0.299 | 1.424 |

**Note:** Subgroup means by C=consensus diagnosis, collapsed across timepoints. Schizoaffective shows the lowest $\text{dAUC}_{\text{norm}}$ and highest $r_{\text{embed}}$ , consistent with more coupled and less divergent conversations than in schizophrenia. Bipolar + psychosis group ( $n = 9$ obs) is underpowered; treat as hypothesis-generating.

### SR10: Hospitalization status

We examined whether hospitalization status (outpatient vs inpatient) moderated the alignment group effect, focusing on dAUC_norm_ and r_embed_ which showed the most prominent diagnostic effects. A PT-only model tested the inpatient–outpatient contrast directly. Within the patient group, no significant inpatient/outpatient difference was observed on either metric, indicating that hospitalisation status does not substantially moderate alignment profiles beyond the broad patient–control distinction.

**Table ST10:**
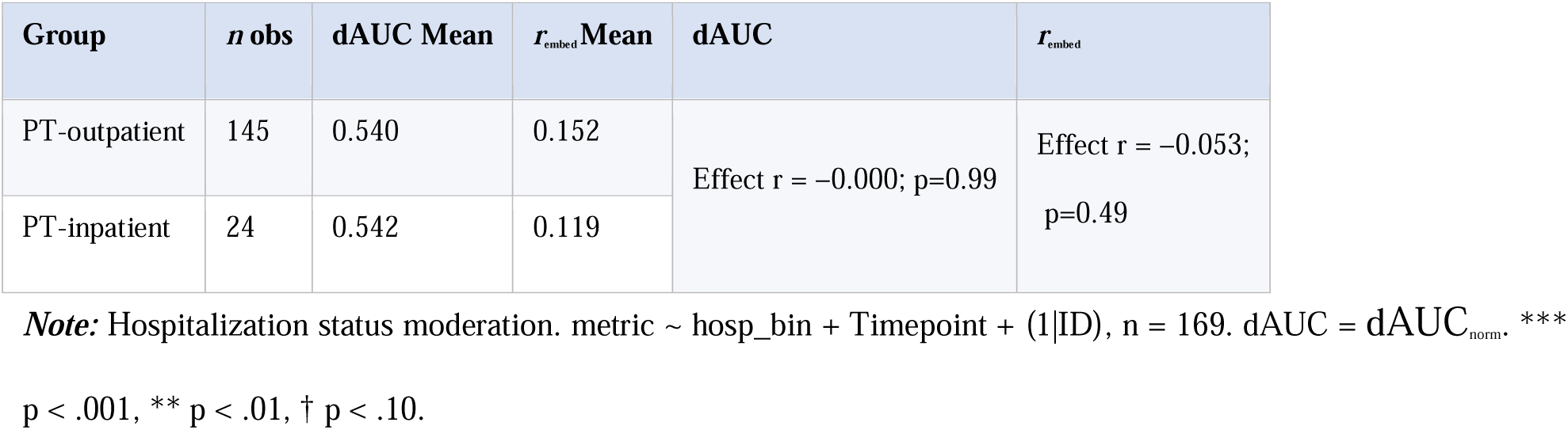
Subgroup Means by Hospitalisation Status.

| Group | n obs | dAUC Mean | $r_{\text{embed}}$ Mean | dAUC | $r_{\text{embed}}$ |
| --- | --- | --- | --- | --- | --- |
| PT-outpatient | 145 | 0.540 | 0.152 | Effect $r = -0.000$; $p=0.99$ | Effect $r = -0.053$;<br>$p=0.49$ |
| PT-inpatient | 24 | 0.542 | 0.119 |  |  |

**Note:** Hospitalization status moderation. $\text{metric} \sim \text{hosp\_bin} + \text{Timepoint} + (1|\text{ID})$ , $n = 169$ . $\text{dAUC} = \text{dAUC}_{\text{norm}}$ . \*\*\*
$p < .001$ , \*\* $p < .01$ , † $p < .10$ .

In addition, both patient subgroups showed elevated dAUC_norm_ relative to HC: the outpatient effect reached significance (r = +0.206, p < .001) and the inpatient effect was a non-significant trend (r = +0.114, p = .060), likely reflecting reduced power.

### SR11. Sensitivity of the principal findings to the first six interviews

Excluding the first six chronologically conducted interviews did not change the direction or nominal statistical interpretation of any pooled patient–control effect. The adjusted patient– control effect for dAUC_norm_ remained positive and statistically supported (full dataset: β = 0.116, p < .001; exclusion sensitivity: β = 0.112, 95% CI [0.066, 0.159], p < .001). The negative patient–control effect for r_embed_ also remained supported (full dataset: β = −0.112, p < .001; exclusion sensitivity: β = −0.109, 95% CI [−0.168, −0.050], p < .001).

Most importantly, the reduction in interviewer within-speaker coherence remained statistically supported and was similar in magnitude after exclusion of the initial interviews (full dataset: β = −0.0151, p = .018; exclusion sensitivity: β = −0.0155, 95% CI [−0.0279, −0.0032], p = .014). The null findings for dAUC_embed_ (β = 0.0449, 95% CI [−0.0476, 0.1374], p = .341) and participant within-speaker coherence (β = 0.0037, 95% CI [−0.0082, 0.0155], p = .546) were likewise unchanged.

No group-by-timepoint interaction was statistically supported after excluding the six interviews: dAUC_norm_, β = −0.0487, p = .298; dAUC_embed_, β = −0.0246, p = .796; r_embed_, β = 0.0243, p = .665; interviewer coherence, β = 0.0140, p = .221; and participant coherence, β = −0.0080, p = .451. Baseline-specific patient–control contrasts also retained their original directions and nominal statistical interpretations.

These findings indicate that the principal results, including reduced interviewer within-speaker coherence in patient interviews, were not materially dependent on the initial six interviews. We therefore retained all interviews in the primary analyses.

### SR12. Speaker-specific semantic trajectories in embedding space

In response to a reviewer’s query on which of the two speakers in a dyad is adapting to the other, we examined each speaker’s semantic trajectory directly. All 271 conversations were encoded with paraphrase-multilingual-MiniLM-L12-v2 (512-token limit). For every conversation, a joint PCA was fitted over both speakers’ paired turn embeddings and each speaker’s PC1 score was followed across the interview (scores z-scored within conversation; 9,055 paired turns).

First, the interviewer–participant PC1 gap showed the same temporal profile as register divergence: the patient–control difference was concentrated early in the interview and dissipated by its end (gap ~ position × group + timepoint + (1 | participant); group β = +0.12, p = .006; position × group β = −0.21, p = .002). Second, anchor-based movement analyses showed that neither speaker moved toward the partner’s early-interview position in either group (all four speaker-by-group cells negative, all p < .0001; role × group p = .07): the temporal attenuation of divergence reflects a change in dyadic distance, not convergence by the participant or accommodation by the interviewer.

Third, turn-level cross-lagged models revealed an asymmetry. In control interviews, semantic following was bidirectional: each speaker’s current turn tracked the partner’s preceding turn (interviewer follows participant, β = +0.090, p < .0001; participant follows interviewer, β = +0.062, p = .004). In patient interviews, the interviewer’s tracking of the participant was selectively attenuated (interaction β = −0.057, p = .009), while participants’ tracking of the interviewer was unchanged (p = .55). Per-conversation refits gave the same pattern (interviewer-follows-participant: HC +0.260 vs PT +0.137, group partial r = −.19, p = .002; participant-follows-interviewer: p = .64). These are observational, lagged associations; they may reflect differential prompting, or the reduced predictability of patient speech, and do not establish mechanism or causal direction.

**Figure SF10:**
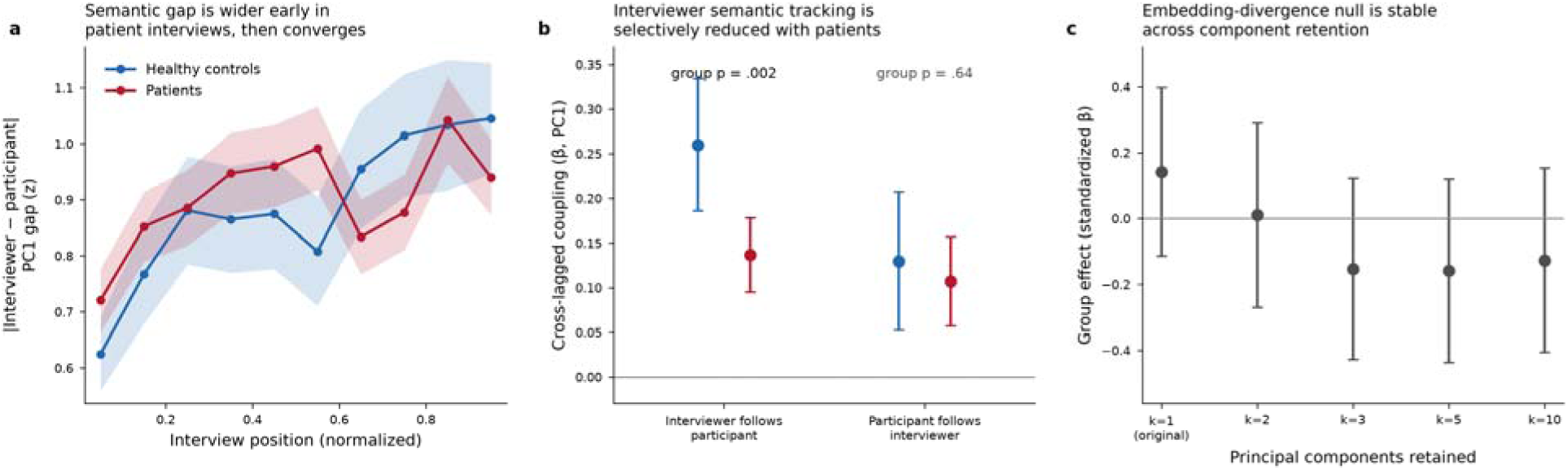
Speaker-specific semantic trajectories in embedding space. ***Note:*** Speaker-specific semantic trajectories in embedding space. (a) Interviewer–participant PC1 gap by normalized interview position (mean ± 95% CI per decile); the patient–control difference is concentrated early. (b) Cross-lagged coupling coefficients (per-conversation fits, mean ± 95% CI): interviewer semantic following of the participant is reduced in patient interviews (p = .002) while participant following of the interviewer is preserved (p = .64). (c) Group effect for embedding-based divergence at 1–10 retained principal components; the null is stable at every component count. Patients in red, healthy controls in blue.

### SR13. Component-retention sensitivity for embedding-based divergence (dAUC_embed_)

dAUCembed was computed by design from the first principal component of a joint within-conversation PCA, which carried a median of 13.0% of within-conversation embedding variance (IQR 11.2–15.4%). Using the turn embeddings, we recomputed dAUCembed retaining 1, 2, 3, 5, and 10 components (Euclidean distance in the retained component space, and variance-weighted composites). The patient–control comparison was null at every component count (standardized β between −0.16 and +0.14, all p > .26; Figure SF10c). The reported null for this metric therefore does not depend on the number of retained components. The primary register-divergence finding (dAUCnorm) involves no PCA.

### SR14. Antipsychotic dose sensitivity

Assessment-day antipsychotic defined daily doses (DDD; WHO ATC/DDD index) were available for 96 of 99 analyzed baseline patients (mean 1.21, SD 0.68) and 58 of 70 patients at 12 months (154 of 169 patient observations). In patient-only models with a participant random intercept, adding standardized DDD left the primary clinical associations unchanged or slightly stronger (register divergence ~ TLI Impoverishment: partial r = +.25, p = .002; synchrony ~ TLI Disorganization: partial r = −.17, p = .032). DDD showed modest independent associations with synchrony (partial r = −.17, p = .034) and interviewer coherence (partial r = −.19, p = .013), reported descriptively; dose likely indexes illness severity in this early-intervention cohort. Antipsychotic exposure therefore does not account for the clinical associations reported in the main text.

## Notes

### Competing Interest Statement

The authors have declared no competing interest.

### Author Declarations

The study was reviewed and approved by the Western University Health Sciences Research Ethics Board.

### Summary of Updates

The manuscript has been revised to clarify the study design, expand the analyses, and qualify the interpretation of the findings. The title and protocol description now explicitly identify the interviews as semi-structured, with standardized tasks and prompts alongside unscripted participant responses and contingent exchanges. New speaker-specific analyses examine semantic trajectories and turn-by-turn associations between interviewer and participant speech. These analyses identify reduced interviewer tracking of participant speech in patient interviews, while participant tracking of interviewer speech is preserved. These findings are presented as observational associations rather than evidence of causal direction or mechanism. Additional sensitivity analyses assess the influence of interviewer identity, extreme thought impoverishment scores, the number of retained principal components in the embedding divergence measure, and antipsychotic dose. Methods and metric definitions have been expanded, and illustrative speech excerpts have been added to aid interpretation. The discussion now distinguishes persistent group differences from exploratory associations with clinical change. Claims concerning interviewer coherence, clinical applications, and generalizability have been qualified. Limitations related to fixed task order, interviewer contributions, conversational structure, and the absence of direct ratings of subjective interpersonal experience are more explicitly acknowledged.

